# Germline Variant Burden Warrants Universal Genetic Testing in Pediatric Myeloid Leukemia

**DOI:** 10.1101/2025.07.29.25332166

**Authors:** Lauren M. Harmon, Zachary S. Hattig, Yizhou Peter Huang, Caliese Beckford, Jason Farrar, Jessica A. Pollard, Sara Zarnegar-Lumley, Xiaotu Ma, Rhonda E. Ries, Soheil Meshinchi, Lucy A. Godley, Timothy J. Triche

**Affiliations:** Van Andel Institute, Grand Rapids, MI, USA; Division of Hematology/Oncology, Department of Medicine, Robert H. Lurie Comprehensive Cancer Center, Northwestern University, Chicago, IL, USA; Michigan State University, East Lansing, MI, USA; Arkansas Children’s Research Institute and University of Arkansas for Medical Sciences, Little Rock, AR, USA; Dana-Farber/Boston Children’s Cancer and Blood Disorders Center, Boston, MA; Northwestern University Feinberg School of Medicine; Ann & Robert H. Lurie Children’s Hospital of Chicago, Chicago, IL, USA; St. Jude Children’s Research Hospital, Memphis, TN, USA; Translational Science and Therapeutics Division, Fred Hutchinson Cancer Center, Seattle, WA, USA; University of Southern California, Los Angeles, CA, USA

**Keywords:** Germline genetics, predisposition, pediatric acute myeloid leukemia

## Abstract

Causal germline genetic variants are frequently detected in young (under age 40) patients presenting with myelodysplastic syndromes (MDS) or bone marrow failure (BMF), where progression to acute myeloid leukemia (AML) contributes substantially to mortality in these patients. We reasoned that de novo pediatric AML, which shares clinical and biological characteristics, might also share germline genetic risk variants. We investigated germline variants in a large cohort (n=365) of pediatric AML patients with whole-genome sequencing (WGS), 29 with matched marrow-derived stromal cells, and 336 with matched remission marrow samples. Variants were deemed “likely germline” based on variant allele frequency (VAF) across available samples. Following American College of Medical Genetics and Genomics (ACMG) and Association of Molecular Pathology (AMP) guidelines, we annotated pathogenic/likely pathogenic (P/LP) variants in 555 genes linked to leukemia risk. P/LP variants were identified in 5.5% (95% CI: (3.3%,7.9%)) of patients in genes linked to familial myeloid malignancy and an additional 3.3% (95% CI: (1.6%,5.2%)) of patients in genes conferring risk to lymphoid malignancy or solid tumors. The large cohort enabled burden testing, which we employed by comparing loss-of-function variants between patients and 2504 control subjects from the 1000 Genomes Project. There was a 6.9-fold (95% CI: (3.1,14.9)) increase in loss-of-function variants in genes implicated in myeloid malignancy risk, a 2.4-fold (95% CI: (1.7,3.2)) increase in candidate risk genes, and a 1.6-fold (95% CI: (1.1,2.3)) increase in randomly-selected genes. We then assembled cohorts totaling 4,622 pediatric and adult patients with acute leukemia or MDS from 10 published studies, and compared P/LP variant burdens across age and diagnosis. The prevalence of germline variants in myeloid malignancies across age groups exceeds 5% consistently and with high confidence. Because the National Comprehensive Cancer Network recommends that all patients receive screening if their pre-test germline variant probability exceeds 5%, our results support germline genetic variant testing as an integral component of diagnostic work-up for myeloid malignancies, including donor selection for stem cell transplantation.

## INTRODUCTION

Leukemia is the most common cancer in young people, and acute myeloid leukemia (AML) increasingly accounts for the bulk of leukemic mortality in this group^1^. Germline genetics have been extensively studied in myelodysplastic syndrome (MDS), which can develop from inherited bone marrow failure (BMF) or other germline predisposition syndromes. Young people (i.e., those under age 40) with bone marrow failure harbor causal germline variants in up to 48% of cases^2^, and causal germline variants have been identified in up to 19% of young MDS patients^3,4^. MDS and AML share similar biological and clinical characteristics, with blast count being the main differentiating feature. However, it is unknown whether germline variants have a similar prevalence among young people with *de novo* AML as seen in MDS. The robust evidence for causal germline risk in BMF and MDS motivated our study of germline variants in a large cohort of young patients with *de novo* AML.

Germline variants in MDS/BMF tend to affect hematopoietic transcription factors, telomere maintenance, DNA repair, or ribosome function. Hematopoiesis requires a high volume of cellular turnover. Germline disruptions that limit replicative potential yield pernicious cytopenias, which can drive selection for cells that accumulate further somatic mutations. For example, biallelic loss-of-function variants in the *SBDS* gene cause Shwachman-Diamond syndrome. Somatic variants in *TP53*^5^ are observed at leukemic progression in SBDS patients, while somatic *EIF6* variants rescue the underlying ribosomal defect without transformation^6^. Germline variants can also contribute to cancer risk via increased genomic instability, increased proliferative capacity, or impaired immune surveillance. Variants in DNA repair pathways can increase the rate of mutation acquisition and clonal evolution, while variants in genes that modulate the RAS pathway, such as *PTPN11* and *NF1,* can accelerate clonal succession. While germline predisposition variants rarely exhibit complete penetrance in childhood, when combined with acquired somatic mutations their impact is often significant^7^.

Germline risk loci for AML have been previously documented, but it is unlikely that all such loci have been identified. A study of seven infants with *KMT2A-*rearranged AML and no known germline predisposition found 1.4 fold more congenital deleterious variants in 655 AML-related genes than in controls (p < 0.001)^8^.

Here we address two primary questions. First, to what extent are known risk alleles for MDS and BMF identified in pediatric *de novo* AML patients? Second, is there an increased burden of deleterious germline variants in candidate risk genes, and if so, which genes account for this difference?

## METHODS

### Overview of cohort characteristics

Pediatric, adolescent, and young adult (ages 0-30 years) patients with AML (N=365) were consented for whole-genome sequencing as part of three Children’s Oncology Group (COG) clinical trials. COG-AAML1031 was the largest single source of probands, with whole genome sequencing (WGS) performed on bone marrow aspirates of 336 patients (139 with samples at diagnosis, remission, and relapse; 117 at diagnosis and remission; 80 at remission and relapse). The NCI/COG TARGET AML Induction Failure cohort, TARGET-21, included 29 patients from two COG clinical trials (COG-AAML03P1 and COG-AAML0531). Due to the presence of malignant cells in all hematopoietic tissues, germline WGS for TARGET-21 was performed on matched bone marrow-derived stromal cells^9^. Matched RNA sequencing data from bone marrow and/or peripheral blood were also available for 358 out of 365 patients (98%). Comprehensive clinical data, including cytogenetic analysis, molecular profiling, outcomes, sex, and age were available for 364 out of 365 participants (99.7%) (Supplementary Table 1, Supplementary Figure 1). This analysis was determined to be non-human subjects exempt by Van Andel Institute’s Institutional Review Board (IRB) based on the universal deidentification of subjects from COG trials (IRB #17027).

### Target gene selection

We examined the variants called in the coding exons, promoters, introns, and 5’ and 3’ untranslated regions of 555 genes associated with hematopoietic malignancies, cancer risk, immunodeficiency, and bone marrow and blood diseases according to prior literature^10,11^ (Supplementary Table 2).

### Assessing likely germline variants and inclusion criteria of variants

Likely germline variants were identified based on variant allele frequency (VAF) >0.3 in bone marrow stromal cell samples. For patients without such samples, we required a VAF >0.3 in all available WGS bone marrow samples (diagnosis, remission, and/or relapse). To minimize sequencing artifacts, variants were filtered for depth >10. To address possible tumor-in-normal contamination or sample swaps, matched remission and tumor WGS samples were manually inspected for presence of somatic driver mutations to confirm that somatic mutations present in the tumor WGS sample were absent, or at a low frequency, in the remission WGS sample. To prevent over-filtering of likely germline variants, including cases of somatic reversion, variants with a VAF>0.3 in the remission sample but with a VAF<0.3 at diagnosis or relapse were manually evaluated for likely germline status based on known germline or somatic role of the gene, frequency of the variant in germline (ClinVar and GnomAD) or somatic (COSMIC) databases, and presence of minimal residual disease in the sample.

### Variant annotation of pathogenicity

We applied the decision tree in Supplementary Figure 2 to prioritize variants for curation programmatically. Variants prioritized included stop gain, frameshift, splice, and start or stop loss variants. Missense variants were prioritized based on REVEL^12^ pathogenicity predictions (score >= 0.7) with an associated PubMed reference as annotated by Variant Effect Predictor^13^, as well as missense variants present in at least two patients and absent in all gnomAD^14^ populations. All variants with a previous pathogenic (P)/likely pathogenic (LP) annotation in ClinVar^15^ were hand curated. All variants were independently evaluated by two biocurators according to American College of Medical Genetics and Genomics/Association of Molecular Pathology guidelines^16^. Where available, modified guidelines for individual genes were used as defined by a Variant Curation Expert Panel (VCEP). If VCEP guidelines were unavailable for a gene, we followed the suggested guidelines for ACMG annotations in hematopoietic malignancies as described^11^. Additional tools used to assist in annotations included VarSome^17^, InterVar^18^, subRVIS^19^, and the UCSC Genome Browser^20^. Variants predicted to impact splicing by the Variant Effect Predictor and SpliceAI^21^ were assessed for splicing abnormalities using matched transcriptomic marrow samples. Genes containing predicted splice variants were visualized for intron retention or exon skipping using Integrated Genome Viewer^22^. Splicing aberrations were applied to annotations as recommended by the ClinGen SVI Splicing Subgroup^23^.

### Statistical Methods

All statistical analyses were performed using R version 4.3.2. Figures were generated using ggplot2^24^. All supporting code is deposited in Zenodo (<u>DOI: 10.5281/zenodo.15586741</u>). Empirical estimates of parameters and test statistics are used wherever possible and provided throughout.

## RESULTS

### Prevalence of P/LP variants in genes associated with myeloid malignancy risk

We first identified P/LP likely germline variants in genes known to confer risk to pediatric and adult myeloid malignancies (Table 1). Six P/LP likely germline variants in pediatric myeloid malignancy risk genes were found in four patients (1.1%). These included *GATA2, ZCCHC8, SBDS, TP53,* and *DNMT3A.* One patient had a heterozygous stopgain variant in *GATA2,* which is associated with *GATA2* Deficiency Disorder, an immune system disorder that increases myeloid malignancy risk^25^. One patient had a splice variant in *ZCCHC8,* and heterozygous loss of function variants in *ZCCHC8* are associated with telomere-related bone marrow failure^26^. A patient had *SBDS* splice variants confirmed to be in the compound heterozygous state, which is associated with Shwachman-Diamond syndrome, a bone marrow failure syndrome characterized by a defect in ribosome biogenesis, resulting in decreased proliferation capability and cytopenias that create a selective advantage for somatic mutations that increase proliferation^5,27^. This patient also had *TP53* p.(Arg248Gln), which is associated with Li-Fraumeni Syndrome, an AML and general tumor predisposition syndrome. However, these mutations have been described as secondary in Shwachman-Diamond syndrome during progression to AML^28^, suggesting that this variant may have been a clonal somatic variant rather than a germline variant. Another patient had *DNMT3A* p.(Arg882His) which was identified in bone marrow stromal cells and therefore unlikely to be the result of tumor contamination. This variant is commonly mutated somatically in myeloid malignancy but a heterozygous germline variant is associated with Tatton-Brown-Rahman syndrome, an overgrowth/intellectual disability syndrome with an increased risk of developing AML^29^. The disorders associated with these germline variants are generally excluded from *de novo* AML COG trials, and as such, all such cases were undiagnosed and unsuspected at the time of trial enrollment. This suggests that germline genetic testing is critical for not only patients and families, but also for the implementation of clinical trials, as certain germline variants may not be clinically detectable by phenotype alone.

**Table 1:**
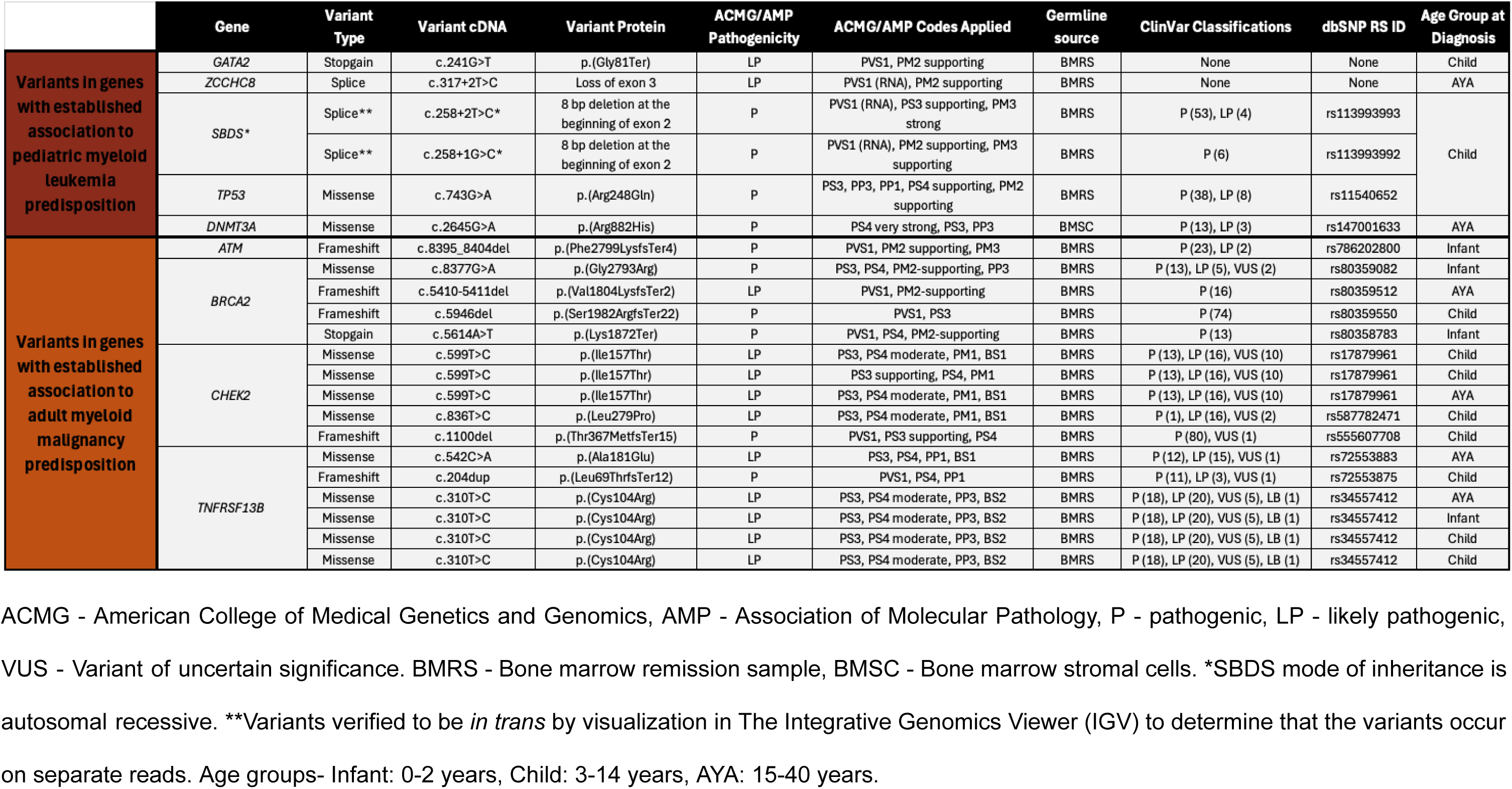
Pathogenic/Likely pathogenic variants in genes associated with myeloid malignancy risk with dominant inheritance.

P/LP likely germline variants in genes that have previously been associated with adult myeloid malignancy were found in 16 patients (4.4%), including *ATM, BRCA2, CHEK2,* and *TNFRSF13B*. No association was found in age at diagnosis and pediatric versus adult predisposition variants. In total, 20 patients (5.5%, 95% CI: (3.3%,7.9%)) had at least one P/LP variant in a gene that has previously been associated with myeloid malignancy risk.

### Prevalence of P/LP variants in genes associated with lymphoid malignancy or solid tumor risk

An additional 13 P/LP variants were found in 12 patients (3.3%) in a gene previously associated with other cancers, including solid tumors (Table 2). These included *HCLS1, BRIP1, PALB2, RAD51C, CDKN2A, MITF, MRE11,* and *SUFU.* In total, 32 patients (8.8%, 95% CI: (6.0%,11.8%)) had a P/LP likely germline variant in a gene associated with myeloid malignancy or cancer risk (Figure 1).

**Figure 1:**
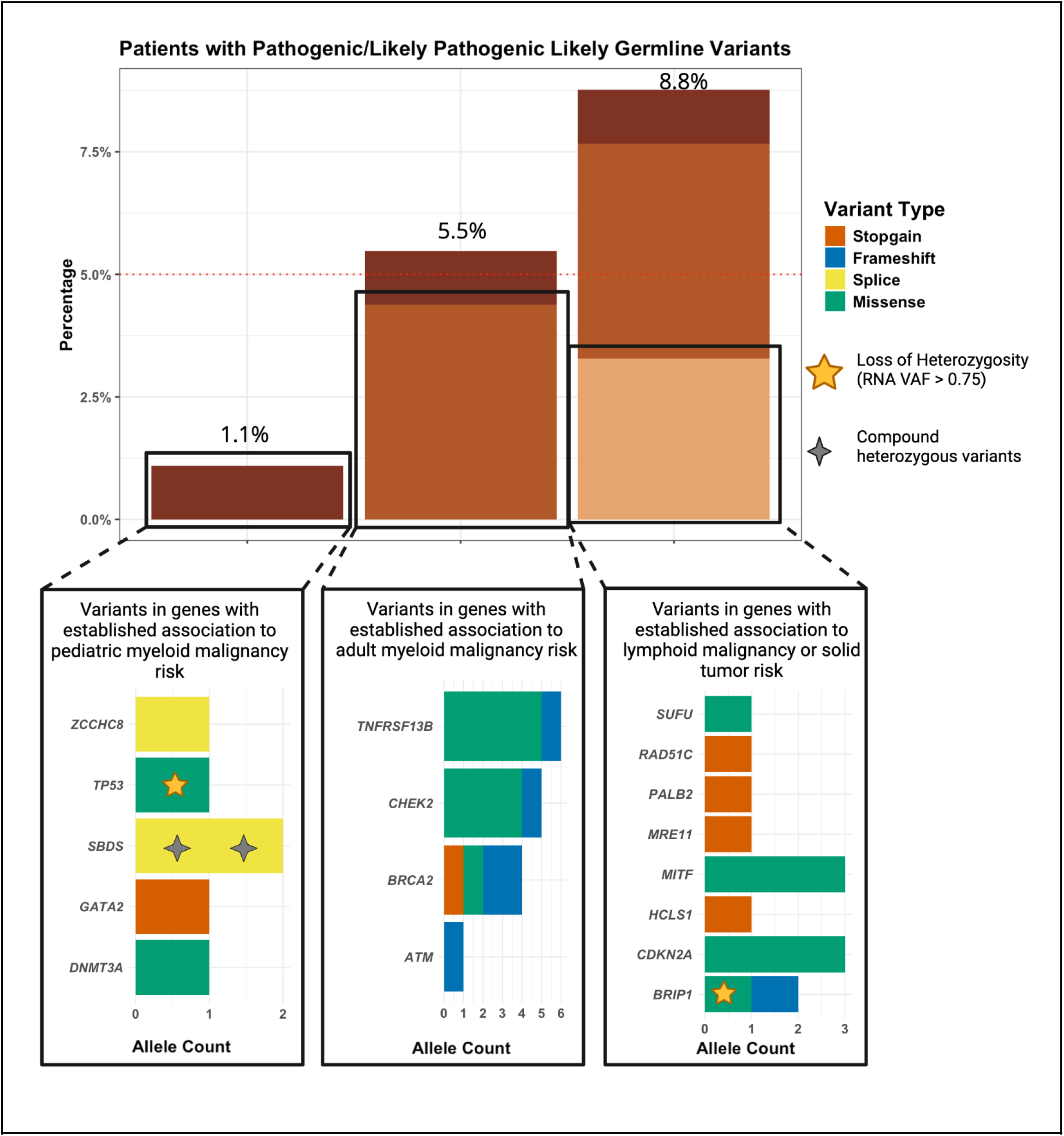
Prevalence of P/LP likely germline variants. Dotted red line represents the 5% threshold defined by the National Comprehensive Cancer Network, which recommends that patients receive screening if their pre-test germline variant probability exceeds 5%^30^.

**Table 2.** Pathogenic/Likely pathogenic variants in genes associated with lymphoid malignancy or solid tumor risk with autosomal dominant inheritance.

| Gene | Variant Type | Variant cDNA | Variant Protein | ACMG/AMP Pathogenicity | ACMG/AMP Codes Applied | Germline source | ClinVar Classifications | dbSNP RS ID | Age Group at Diagnosis |
| --- | --- | --- | --- | --- | --- | --- | --- | --- | --- |
| *HCLS1* | Stopgain | c.1191T>G | p.(Tyr397Ter) | LP | PVS1, PM2 supporting | BMRS | None | rs1383787778 | Child |
| *BRIP1* | Missense | c.139C>G | p.(Pro47Ala) | LP | PS3 moderate, PS4 moderate, PM1, PP3 | BMRS | P (2), VUS (22), LB (6), B (1) | rs28903098 | AYA |
|  | Frameshift | c.2990-2993del | p.(Thr997ArgfsTer61) | P | PVS1, PS4 | BMRS | P (17), LP (9), VUS (1) | rs771028677 | Infant |
| *PALB2* | Stopgain | c.3113G>A | p.(Trp1038Ter) | P | PVS1, PS4 moderate, PP1 | BMRS | P (31), LP (1) | rs180177132 | AYA |
| *RAD51C* | Stopgain | c.577C>T | p.(Arg193Ter) | P | PVS1, PS4 | BMRS | P (17), LP (3) | rs200293302 | Child |
| *CDKN2A* | Missense | c.194T>C | p.(Leu65Pro) | LP | PS4, PP1 moderate, BP4 | BMSC | P (1), LP (3), VUS (1) | rs1587332314 | Infant |
|  | Missense | c.194T>C | p.(Leu65Pro) | LP | PS4, PP1 moderate, BP4 | BMSC | P (1), LP (3), VUS (1) | rs1587332314 | Child |
|  | Missense | c.194T>C | p.(Leu65Pro) | LP | PS4, PP1 moderate, BP4 | BMSC | P (1), LP (3), VUS (1) | rs1587332314 | Infant |
| *MITF* | Missense | c.1273G>A | p.(Glu425Lys) | LP | PS3 moderate, PS4 moderate, PM1, PP1 | BMRS | P (18), LP (7) | rs149617956 | Infant |
|  | Missense | c.1273G>A | p.(Glu425Lys) | LP | PS3 moderate, PS4 moderate, PM1, PP2 | BMRS | P (18), LP (7) | rs149617956 | Child |
|  | Missense | c.1273G>A | p.(Glu425Lys) | LP | PS3 moderate, PS4 moderate, PM1, PP3 | BMRS | P (18), LP (7) | rs149617956 | Child |
| *MRE11* | Stopgain | c.1714C>T | p.(Arg572Ter) | P | PVS1, PS4 | BMRS | P (6), LP (2) | rs137852761 | Infant |
| *SUFU* | Missense | c.367C>T | p.(Arg123Cys) | LP | PP1 Strong, PS3 moderate, PP3, PM2 supporting | BMRS | VUS (2) | rs202247756 | AYA |

ACMG - American College of Medical Genetics and Genomics, AMP - Association of Molecular Pathology, P - pathogenic, LP - likely pathogenic, VUS - Variant of uncertain significance. BMRS - Bone marrow remission sample, BMSC - Bone marrow stromal cells. Age groups- Infant: 0-2 years, Child: 3-14 years, AYA: 15-40 years.

### Prevalence of P/LP variants in genes with limited evidence for association to cancer risk

We identified additional P/LP likely germline variants that at this time are not known to cause cancer risk, but may warrant future investigation. These variants, with brief summaries of the literature supporting germline risk for each gene are included in Supplementary Tables 3-9. P/LP variants found in genes with moderate evidence for association to hematopoietic malignancy risk in the heterozygous form included *ERBB2, CASP10, DHX34* (n=2), *PRF1* (n=4), *FANCA*, and *RASA2* (Supplementary Table 3). P/LP variants in genes with limited evidence for association to hematopoietic malignancy risk were found in *DDX54, DNAH5* (n=4)*, DNAH9* (n=2), and *ATG2B* (Supplementary Table 4). P/LP variants with limited evidence for association to solid tumor risk in the heterozygous form were found in *SMO* (n=2)*, ROS1* (n=2), *FANCM, SLX4* (n=2), and *RECQL5* (Supplementary Table 5). In total, 22 patients (6.0%, 95% CI: (3.8%,8.5%)) had at least one P/LP variant in a gene with limited or moderate evidence for association to cancer risk.

### Heterozygous P/LP variants in genes with autosomal recessive disease inheritance

Heterozygous P/LP variants in genes with autosomal recessive hematopoietic malignancy risk inheritance were identified in *CTC1* (n=2)*, FANCD2, RAD50, RPS27A, STXBP2, VPS45,* and *XRCC2* (Supplementary Table 6). Heterozygous P/LP variants in genes with autosomal recessive solid tumor risk inheritance included *MUTYH* (n=5), *NTHL1, RECQL4,* and *XPC* (Supplementary Table 7). Heterozygous P/LP variants in genes associated with other hematopoietic diseases or developmental disorders with autosomal recessive inheritance were identified in *ADAMTS13, GBA1* (n=3), and *VPS13B* (Supplementary Table 8). Although there is insufficient evidence to conclude that these variants contribute to disease risk, future research may clarify the extent to which dosage in these genes affects disease risk.

### P/LP variants in genes with a known somatic role in AML but no prior evidence for germline malignancy risk

P/LP likely germline variants were also identified in genes that are known to be somatically mutated in AML but have no evidence in the literature for conferring germline malignancy risk (Supplementary Table 9). These included *ARHGEF12, BRCC3, BRD4, CUX1, FGFR3, INPP5D, NFKBIE, PRPF8* (n=2)*, SETDB1, SYNE1,* and *MYO5A.* Future research is needed to determine the effects of these variants for conferring germline risk.

### Characteristics of patients with P/LP likely germline variants

Patients harboring a P/LP germline variant did not show significant differences in the clinical characteristics or demographics of sex, age, ethnicity, AML somatic driver subtype, or event-free survival (Supplementary Figure 3). The co-occurrence of P/LP germline and somatic variants is summarized in Supplementary Figure 4. Patients with multiple P/LP variants are reported in Supplementary Table 10. Patient PAUVWV carried P/LP variants in *MRE11A* and *MUTYH*, both vital to DNA repair. Additional characteristics of patients with P/LP variants are included in Supplementary Table 11, including variant allele frequencies of each variant in each sample, tumor-in-normal estimates, minimal residual disease of the sample, as well as the sex, age, ethnicity, AML subtype, and clinical outcome for each patient.

### Analysis of second hits in genes with P/LP variants

Second hits in genes with P/LP variants provide additional evidence for causality, especially when they occur in genes which confer increased disease risk when both copies are affected. To determine if patients with P/LP variants also had a second hit in the same gene, we analyzed loss of heterozygosity (LOH) and additional somatic variants.

LOH occurs when only the variant allele is expressed in the RNA. This can occur through varied mechanisms such as deletion or epigenetic silencing of the wild-type allele. To identify LOH, we compared the VAF for each P/LP variant in genomic and transcriptomic diagnosis samples (Supplementary Figure 5). LOH occurred in P/LP variants in *BRIP1* p.(Pro47Ala) and *TP53* p.(Arg248Gln), as evidenced by transcriptomic VAFs nearing 100%, indicating that LOH led to loss of expression of the wild-type allele*. BRIP1* is associated with solid tumor risk when one copy is affected, and Fanconi anemia when both copies are affected, which leads to bone marrow failure and leukemia risk. The LOH in the *BRIP1* p.(Pro47Ala) variant provides evidence of causality for this patient’s leukemia. *TP53* variants are associated with compensatory clonal hematopoiesis in patients with Shwachman-Diamond disorder by enhancing clone fitness. Accordingly, the patient with *TP53* p.(Arg248Gln) displayed a somatic deletion of a region of chromosome 17 including the *TP53* gene, explaining the increase in transcriptomic VAF of the variant.

Similarly, secondary loss-of-function somatic variants in genes with a P/LP variant can also provide evidence for causality. Patients harboring P/LP variants were scanned for additional somatic variants occurring in the same gene. Secondary deleterious somatic variants included a splice variant in *FGFR3,* a missense and a splice variant in *PRPF8,* and a missense variant in *CUX1* (Supplementary Table 12). These three genes are not known to contribute to germline risk, but have been documented as loci with recurrent somatic alterations in AML^31–33^. More research is needed to determine the germline role of these genes in AML risk.

### Burden testing comparisons with healthy controls

We tested if pediatric AML patients have a higher burden of deleterious variants than the general population. We compared them to 2,504 healthy controls from the 1000 Genomes Project^34^. Using the Ensembl Variant Effect Predictor^13^, we identified putative loss-of-function (pLoF) variants caused by a stop gain or a frameshift affecting the primary gene transcript and with a frequency less than 0.001 in GnomAD^14^. To estimate the uncertainty of our estimates, we performed permutation testing by taking resamples of 90% of each group of genes. In genes with an established role in familial myeloid malignancy with dominant inheritance as previously reviewed^35,36^, 3.9% of pediatric AML patients had a pLoF variant, compared to 0.6% of healthy controls (odds ratio = 6.9, 95% CI: (3.1,14.9), p < 0.001) (Figure 2A). To determine if there is also an increased burden of deleterious variants in candidate genes for leukemia risk, we examined pLoF variants in 484 additional genes, and found that 14.0% of pediatric AML patients and 6.4% of 1000 genomes subjects had a pLoF variant in these genes (odds ratio = 2.4, 95% CI: (1.7,3.2), p < 0.001), suggesting that variants in these genes also contribute to leukemia risk. For comparison, we also analyzed 500 random genes. Here, 9.2% of pediatric AML patients and 5.7% of healthy controls had a pLoF variant (odds ratio = 1.6, 95% CI: (1.1,2.3), p = 0.01). This indicates that AML patients overall may harbor an increased burden of deleterious variants, and this difference is more pronounced in genes thought to contribute to leukemia risk. To mitigate the possibility of bias due to differences in overall number of variants, we examined the overall number of variants in these genes, finding a slight increase in number of variants in control subjects as compared to pediatric AML patients, which suggests that these odds ratios are likely an underestimate (Supplementary Figure 6). In genes in the homologous recombination repair pathway that have previously been shown to confer breast cancer risk in autosomal dominant form, 0.08% of healthy controls and 1.8% of pediatric AML patients had a pLoF variant (odds ratio = 22.7, 95% CI: (5.2,155.5), p < 0.001).

**Figure 2:**
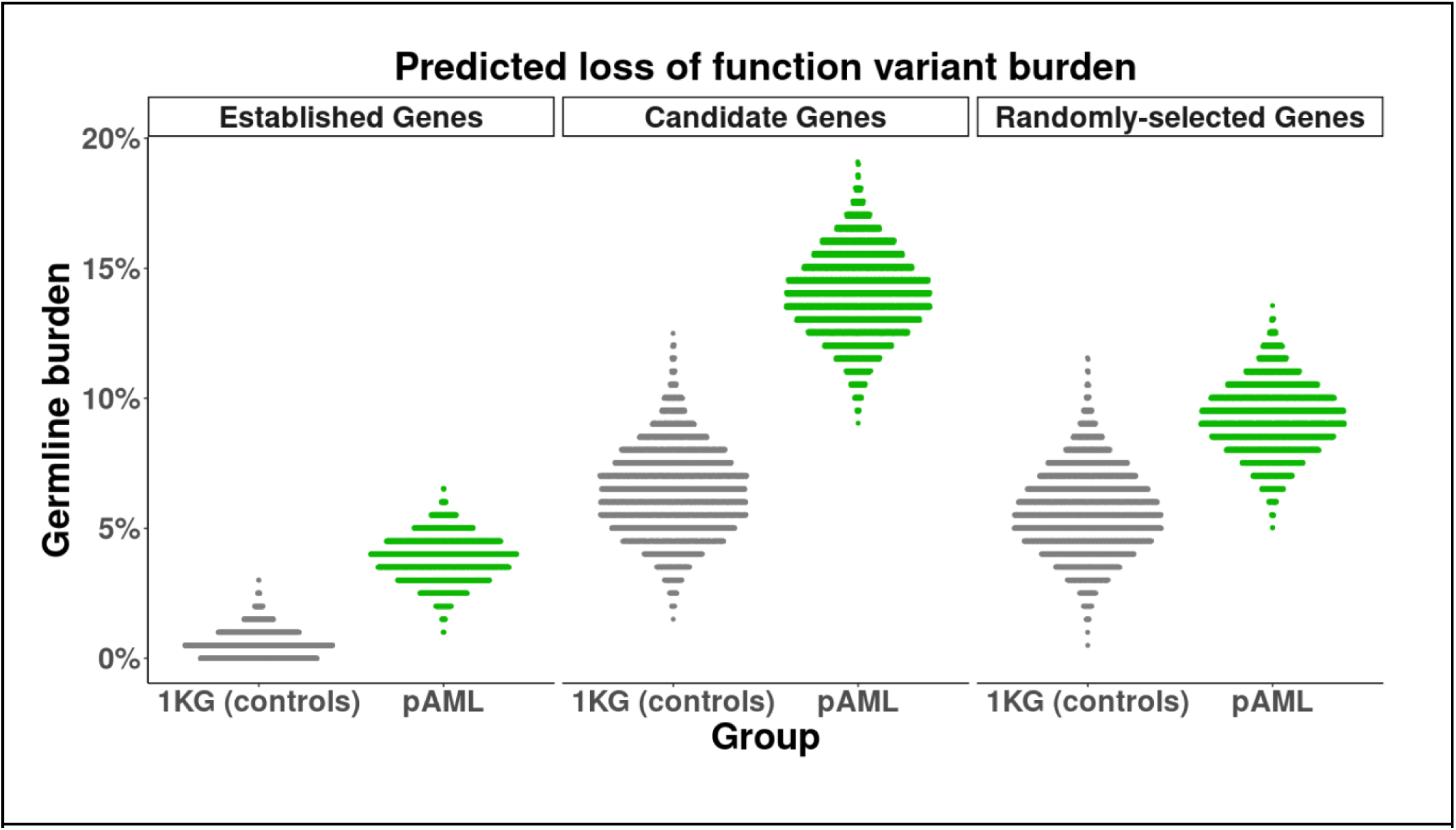
Comparisons of germline burden in 1000 Genomes Project control subjects. a) Percentage of subjects with a predicted loss-of-function variant, using subsamples of all subjects. Established genes include genes with a known association to myeloid malignancy risk. Candidate genes are selected genes for possible myeloid malignancy risk association. 1KG - 1000 Genomes Project.

### Meta-analysis of germline burden in pediatric and adult ALL, AML, and MDS

We compared the overall prevalence of P/LP likely germline variants in pediatric AML to that of acute lymphoblastic leukemia (ALL) and MDS. We analyzed 10 published studies^3,4,10,11,37–42^, involving 4,622 patients with germline variant annotations (Figure 3A). To control for differences in gene panels, we included only variants in the genes from the University of Chicago’s hereditary hematopoietic malignancies/immunodeficiencies gene panel (Supplementary Table 13). P/LP variants were required to be homozygous or compound heterozygous for genes with autosomal recessive modes of inheritance. A robust Poisson test for trend showed germline burdens significantly higher (P<0.001) in AML (6.5%, 95% CI: (4.9%,8.4%), N=859) than in B-cell ALL (1.8%, 95% CI: (1.4%, 2.4%), N=2560) or T-cell ALL (2.1%, 95% CI: (1.1%, 3.7%), N=566), but lower than in MDS (14.1%, 95% CI: (11.5%, 17.0%), N=651). We performed resampling to estimate the uncertainty by taking 250 random samples from each group, employing 3,333 resamples for each group. ALL, AML, and MDS show distinct distributions of germline burden (Figure 3B).

**Figure 3:**
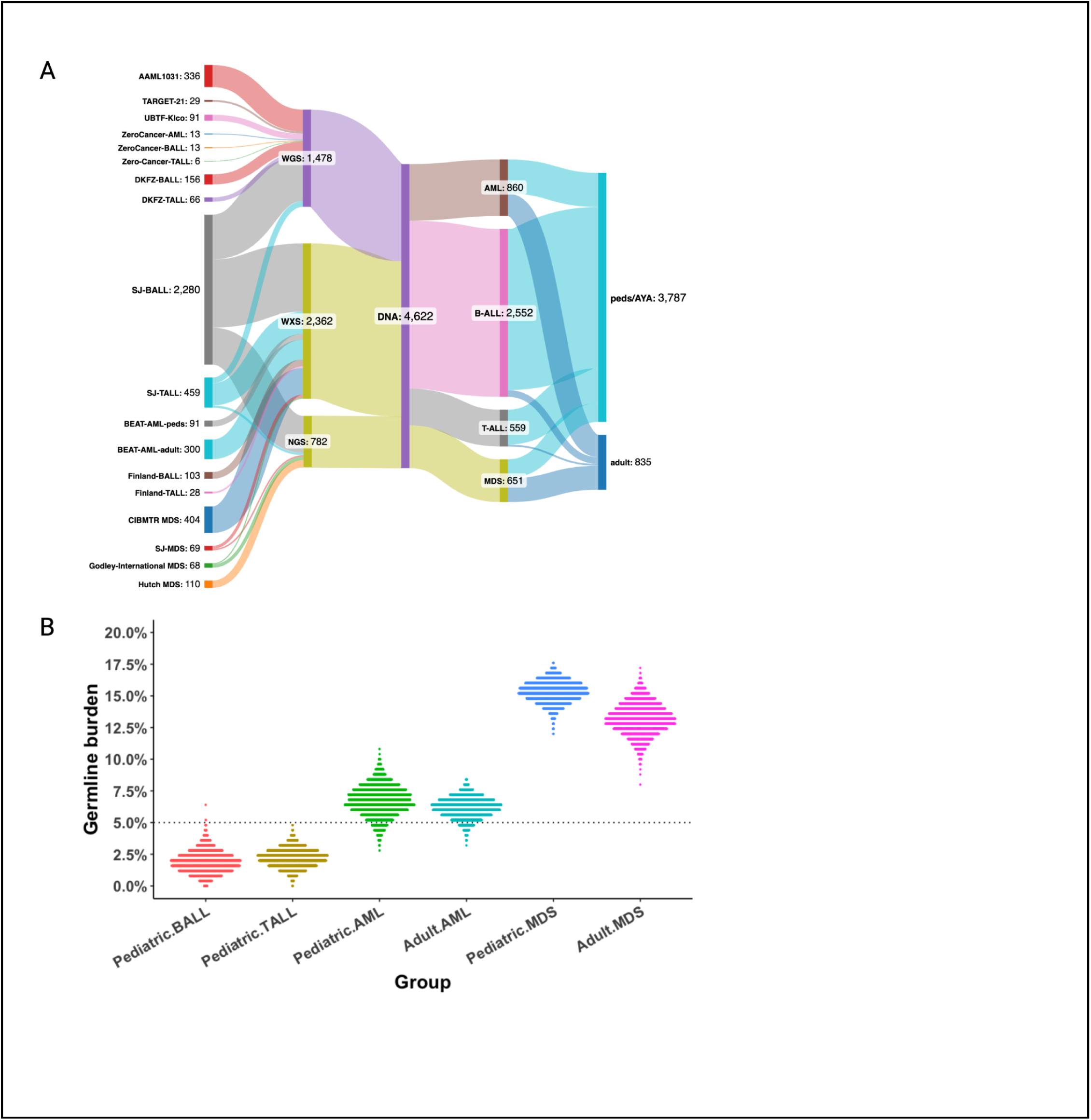
Comparisons of germline burden in pediatric and adult AML, ALL, and MDS. a) Flow diagram of datasets included, created with SankeyMATIC. b) Distributions of germline burden in resampled cohorts. ALL - acute lymphoblastic leukemia. AML - acute myeloid leukemia. MDS - myelodysplastic syndrome. WGS - whole genome sequencing. WXS - whole exome sequencing. NGS - next generation sequencing. AYA - adolescent young adults.

## DISCUSSION

This study represents the largest study to date of germline genetic variants in pediatric AML patients. The prevalence of deleterious germline variants is high in this cohort of patients with presumed *de novo* AML. Using conservative manual variant annotation according to ACMG/AMP guidelines, we identified 5.5% (95% CI: (3.3%,7.9%)) of patients with at least one P/LP variant in genes with established evidence for AML risk, and 8.8% (95% CI: (6.0%,11.8%)) of patients with at least one P/LP variant including genes with established associations to lymphoid or solid tumor risk. According to The National Comprehensive Cancer Network, germline screening is recommended if the pre-test probability is >5%^30^. Therefore, even our most conservative estimate of germline prevalence (5.5%) qualifies pediatric AML patients to receive germline cancer risk testing. This has subsequent implications for future disease surveillance and family screening. It is also of major importance for patients receiving hematopoietic stem cell transplant, since donors are often related family members, and selection of a donor without a germline risk variant should be prioritized.

These results show the potential benefit of integrating germline genetic testing into the diagnostic work-up for pediatric AML. Variants in *GATA2, ZCCHC8, SBDS, TP53,* and *DNMT3A* indicate that some unrecognized inherited BMF or other germline predisposition syndromes still present with AML as the sole finding. These patients would likely benefit from being cataloged and treated according to their specific conditions. For example, we identified a patient with biallelic variants in *SBDS,* which is associated with Shwachman-Diamond syndrome (SDS). This patient also had a *TP53* heterozygous variant that exhibited loss of heterozygosity in the tumor sample. Patients with SDS and *TP53* variants have a high rate of clonal hematopoiesis^43^, and do not tend to respond well to conventional chemotherapy treatment^44^. Accordingly, this patient was positive for minimal residual disease after both cycles of induction therapy. However, patients with germline or somatic *TP53* variants have shown some response to hypomethylating agents^45^. Germline genetic characterization for patients such as this one would provide crucial information for identifying an appropriate treatment plan. Furthermore, whole genome sequencing can be done in a clinically relevant timeframe and can lead to improved outcomes^46^.

Our results show that genes that have previously been associated with adult myeloid malignancy also affect pediatric patients. *CHEK2* has been thoroughly reviewed for its role in myeloid malignancy risk^47^. Its odds ratio for MDS is 7.6^48^. The I157T variant, while relatively common, has also been documented to carry an odds ratio of 6.44 for hematopoietic malignancies, indicating a pathogenic role with low penetrance^49^. A mouse model of *Chek2* I161T (homologous to *CHEK2* I157T) had fewer Lin-CD34+ and Lin-cKit+ cells and inferior survival which was likely due to development of hematological malignancies^49^. *ATM* germline variants have been shown to increase hematopoietic malignancy risk in adults, and the odds ratio for chronic lymphocytic leukemia is 14.83^47^. *ATM* nonsense variants were found in 6.4% of adult AML patients^50^ and have also been identified as a strong risk locus for myeloid gene clonal hematopoiesis^51^. *ATM* and *CHEK2* variants create a higher risk of generating chromosomal translocations and other mutations associated with myeloid leukemia development. *ATM* also plays an essential role in HSC reconstitutive capacity^52^.

We identified 4 individuals (1.1%) with P/LP variants in *BRCA2,* which has previously been associated with an increased prevalence in adult myeloid malignancy, where the odds ratio for loss-of-function germline variants was 4.65 (95% CI: (2.29,9.43), P<0.0001)^53^. However, the role of *BRCA1/BRCA2* in *de novo* hematopoietic malignancies is still controversial, with some contending that there is insufficient evidence to establish a causal role^54^. Our results provide additional evidence in support of the role of germline *BRCA2* in AML risk. Three of the four *BRCA2* cases had a loss-of-function variant (stop gain or frameshift), or 0.8% of the cohort. *BRCA2* loss-of-function variants are present in 0.14%-0.22% of the general population^30,55^; therefore the incidence in pediatric AML is 4.57 times higher than the general population, which is concordant with the reported odds ratio from Stubbins et al^53^.

Similarly, P/LP variants in *TNFRSF13B* found in 6 patients (1.6%) provide strong support for the inclusion of *TNFRSF13B* variants in germline screening for pediatric AML patients. *TNFRSF13B* variants have been reported in patients with MDS^11^, Hodgkin lymphoma^56^, and have been shown to be associated with chronic lymphocytic leukemia risk^57^ and multiple myeloma risk^58^. The P/LP variants *TNFRSF13B* C104R (n=4) and A181E (n=1) are common in patients with common variable immunodeficiency, which is known to increase hematopoietic malignancy risk^59^. C104R has been documented in a family with AML^60^. C104R and A181E have been shown to abolish TACI signalling^61,62^, and both subsequently impair B-cell function^63^. The ligand for *TNFRSF13B* is *TNFSF13B,* which has been shown to be a positive regulator of AML-initiating cells by reducing apoptosis and increasing proliferation^64^. This evidence suggests that *TNFRSF13B* should be considered a risk locus for AML with low penetrance.

The prevalence of P/LP variants in genes known to increase risk for lymphoid malignancy or solid tumors is high (3.3%). Therefore, further study of the role of these genes in AML risk should be a priority for future research. For example, three patients had P/LP variants in *CDKN2A,* which is well-known for melanoma and other cancer risk^65^. *CDKN2A* deletion has also been associated with poor outcomes in B-cell leukemias^66,67^. All three of these patients failed induction therapy, and one patient had a somatic loss of chromosome 9p. Three patients had the variant *MITF* E419K, which has been associated with a 5 times risk of melanoma, and prevents the MITF protein from being SUMOylated, resulting in impaired cellular senescence^68,69^. *BRIP1* heterozygous variants have been implicated in ovarian cancer risk^70^. We identified two patients with *BRIP1* heterozygous variants, with one of these patients exhibiting loss of heterozygosity in their tumor samples. Future research should focus on the potential role of germline variants in these and other genes associated with solid tumor risk.

Our results indicate that much remains to be discovered in germline risk for pediatric AML. By comparing pediatric AML patients to control subjects from the 1000 Genomes Project, we saw an increase in putative loss-of-function variants not only in genes known to contribute to AML risk, but also in both randomly-selected and candidate genes for AML risk. This indicates that more genes are likely involved in AML risk than are currently appreciated. Many of these variants occurred in genes with limited evidence for malignancy risk, or in genes that are known to occur somatically in AML. The evidence supporting these genes is discussed in Supplementary Tables 3-9. These variants may explain a portion of the missing heritability in pediatric AML, and we suggest that these genes be prioritized for future study.

Finally, the prevalence of P/LP germline variants in a meta-analysis supports the indication that pediatric AML has a higher germline burden than pediatric B-ALL and T-ALL, but a lower burden than pediatric MDS. Although limited by differences in germline testing and analysis methods, this analysis suggests a mechanistic role for interactions of heritable risk and exposures in myeloid malignancy. The structural variants which drive pediatric ALL are attributable, in part, to off-target activity of *AID* and *RAG* genes, which are mandatory for B and T cell maturation^71^. By contrast, our results provide evidence that the likelihood of developing pediatric AML is increased by germline variants. Therefore, germline variants may be considered an integral part of disease etiology and thereby inform more effective surveillance strategies for this rare disease.

Benefits to germline genetic testing in pediatric AML reach beyond disease risk and mechanisms. Stem cell transplantation is warranted in children with high-risk AML, and germline variants can profoundly influence donor selection. Cascade testing of family members can promote surveillance and early detection of malignancies. Finally, modern sequencing platforms have rapid turnaround and plummeting costs and facilitate sequencing of patients and parents at first encounter. We propose that universal germline genetic testing is mandatory to advance care in pediatric AML.

## DATA AVAILABILITY

The data that support the findings of this study are available upon access request from the TARGET: Acute Myeloid Leukemia (AML) project in dbGaP (study accession: phs000465.v23.p8).

## CODE AVAILABILITY

The code used for burden testing and the cohort comparison analysis is deposited in Zenodo (DOI: 10.5281/zenodo.15586741).

## ACKNOWLEDGEMENTS

Computation for the work described in this paper was supported by the High Performance Cluster and Cloud Computing (HPC3) Resource at the Van Andel Research Institute and the Van Andel Institute VAI Bioinformatics and Biostatistics Core Facility (RRID:SCR_024762).

## AUTHOR CONTRIBUTIONS

LH, LG, and TT conceptualized the study. LH designed the computational workflow, performed the bioinformatic and statistical analyses, data interpretations, visualizations, and manuscript drafting. LH and ZH annotated variants according to ACMG/AMP guidelines. YH assisted in implementing the computational pipeline. CB assisted in data visualizations. JF, JP, SZ, and XM assisted in data interpretations. XM performed variant calling from sequencing samples. RR and SM curated and advised on use of sequencing and clinical metadata. LG and TT supervised the project and revised the manuscript. LH wrote and revised the manuscript with input from all authors.

## REFERENCES

1. Siegel RL, Miller KD, Wagle NS, Jemal A. Cancer statistics, 2023. CA Cancer J Clin 2023;73(1):17–48.

2. Bluteau O, Sebert M, Leblanc T, et al. A landscape of germ line mutations in a cohort of inherited bone marrow failure patients. Blood 2018;131(7):717–32.

3. Keel SB, Scott A, Sanchez-Bonilla M, et al. Genetic features of myelodysplastic syndrome and aplastic anemia in pediatric and young adult patients. Haematologica 2016;101(11):1343–50.

4. Feurstein S, Churpek JE, Walsh T, et al. Germline variants drive myelodysplastic syndrome in young adults. Leukemia 2021;35(8):2439–44.

5. Reilly CR, Shimamura A. Predisposition to myeloid malignancies in Shwachman-Diamond syndrome: biological insights and clinical advances. Blood 2023;141(13):1513–23.

6. Tan S, Kermasson L, Hilcenko C, et al. Somatic genetic rescue of a germline ribosome assembly defect. Nat Commun 2021;12(1):5044.

7. Knudson AG Jr. Mutation and cancer: statistical study of retinoblastoma. Proc Natl Acad Sci U S A 1971;68(4):820–3.

8. Valentine MC, Linabery AM, Chasnoff S, et al. Excess congenital non-synonymous variation in leukemia-associated genes in MLL-infant leukemia: a Children’s Oncology Group report. Leukemia 2014;28(6):1235–41.

9. McNeer NA, Philip J, Geiger H, et al. Genetic mechanisms of primary chemotherapy resistance in pediatric acute myeloid leukemia. Leukemia 2019;33(8):1934–43.

10. Yang F, Long N, Anekpuritanang T, et al. Identification and prioritization of myeloid malignancy germline variants in a large cohort of adult patients with AML. Blood 2022;139(8):1208–21.

11. Feurstein S, Trottier AM, Estrada-Merly N, et al. Germ line predisposition variants occur in myelodysplastic syndrome patients of all ages. Blood 2022;140(24):2533–48.

12. Ioannidis NM, Rothstein JH, Pejaver V, et al. REVEL: An Ensemble Method for Predicting the Pathogenicity of Rare Missense Variants. Am J Hum Genet 2016;99(4):877–85.

13. McLaren W, Gil L, Hunt SE, et al. The Ensembl Variant Effect Predictor. Genome Biol 2016;17(1):122.

14. Chen S, Francioli LC, Goodrich JK, et al. A genome-wide mutational constraint map quantified from variation in 76,156 human genomes [Internet]. bioRxiv. 2022 [cited 2023 Nov 21];2022.03.20.485034. Available from: https://www.biorxiv.org/content/10.1101/2022.03.20.485034v2

15. Landrum MJ, Lee JM, Benson M, et al. ClinVar: improving access to variant interpretations and supporting evidence. Nucleic Acids Res 2018;46(D1):D1062–7.

16. Richards S, Aziz N, Bale S, et al. Standards and guidelines for the interpretation of sequence variants: a joint consensus recommendation of the American College of Medical Genetics and Genomics and the Association for Molecular Pathology. Genet Med 2015;17(5):405–24.

17. Kopanos C, Tsiolkas V, Kouris A, et al. VarSome: the human genomic variant search engine. Bioinformatics 2019;35(11):1978–80.

18. Li Q, Wang K. InterVar: Clinical Interpretation of Genetic Variants by the 2015 ACMG-AMP Guidelines. Am J Hum Genet 2017;100(2):267–80.

19. Gussow AB, Petrovski S, Wang Q, Allen AS, Goldstein DB. The intolerance to functional genetic variation of protein domains predicts the localization of pathogenic mutations within genes. Genome Biol 2016;17:9.

20. Kent WJ, Sugnet CW, Furey TS, et al. The human genome browser at UCSC. Genome Res 2002;12(6):996–1006.

21. Jaganathan K, Kyriazopoulou Panagiotopoulou S, McRae JF, et al. Predicting Splicing from Primary Sequence with Deep Learning. Cell 2019;176(3):535–48.e24.

22. Robinson JT, Thorvaldsdóttir H, Wenger AM, Zehir A, Mesirov JP. Variant Review with the Integrative Genomics Viewer. Cancer Res 2017;77(21):e31–4.

23. Walker LC, Hoya M de la, Wiggins GAR, et al. Using the ACMG/AMP framework to capture evidence related to predicted and observed impact on splicing: Recommendations from the ClinGen SVI Splicing Subgroup. Am J Hum Genet 2023;110(7):1046–67.

24. Wickham H. ggplot2: Elegant Graphics for Data Analysis. Springer Science & Business Media; 2009.

25. Hsu AP, McReynolds LJ, Holland SM. GATA2 deficiency. Curr Opin Allergy Clin Immunol 2015;15(1):104–9.

26. Savage SA, Niewisch MR. Dyskeratosis congenita and related telomere biology disorders. In: GeneReviews(®). Seattle (WA): University of Washington, Seattle; 1993.

27. Nelson A, Myers K. Shwachman-Diamond syndrome. In: GeneReviews(®). Seattle (WA): University of Washington, Seattle; 1993.

28. Kennedy AL, Myers KC, Bowman J, et al. Distinct genetic pathways define pre-malignant versus compensatory clonal hematopoiesis in Shwachman-Diamond syndrome. Nat Commun 2021;12(1):1334.

29. Ostrowski PJ, Tatton-Brown K. Tatton-Brown-Rahman syndrome. In: GeneReviews(®). Seattle (WA): University of Washington, Seattle; 1993.

30. Daly MB, Pilarski R, Yurgelun MB, et al. NCCN Guidelines Insights: Genetic/Familial High-Risk Assessment: Breast, ovarian, and pancreatic, version 1.2020. J Natl Compr Canc Netw 2020;18(4):380–91.

31. Guo C, Ran Q, Sun C, et al. Loss of FGFR3 delays acute myeloid leukemogenesis by programming weakly pathogenic CD117-positive leukemia stem-like cells. Front Pharmacol 2020;11:632809.

32. Kurtovic-Kozaric A, Przychodzen B, Singh J, et al. PRPF8 defects cause missplicing in myeloid malignancies. Leukemia 2015;29(1):126–36.

33. McNerney ME, Brown CD, Wang X, et al. CUX1 is a haploinsufficient tumor suppressor gene on chromosome 7 frequently inactivated in acute myeloid leukemia. Blood 2013;121(6):975–83.

34. 1000 Genomes Project Consortium, Auton A, Brooks LD, et al. A global reference for human genetic variation. Nature 2015;526(7571):68–74.

35. Rafei H, DiNardo CD. Hereditary myeloid malignancies. Best Pract Res Clin Haematol 2019;32(2):163–76.

36. University of Chicago Hematopoietic Malignancies Cancer Risk Team. How I diagnose and manage individuals at risk for inherited myeloid malignancies. Blood 2016;128(14):1800–13.

37. Douglas SPM, Lahtinen AK, Koski JR, et al. Enrichment of cancer-predisposing germline variants in adult and pediatric patients with acute lymphoblastic leukemia. Sci Rep 2022;12(1):10670.

38. Junk SV, Förster A, Schmidt G, et al. Germline variants in patients developing second malignant neoplasms after therapy for pediatric acute lymphoblastic leukemia-a case-control study. Leukemia [Internet] 2024;Available from: 10.1038/s41375-024-02173-2

39. Brady SW, Roberts KG, Gu Z, et al. The genomic landscape of pediatric acute lymphoblastic leukemia. Nat Genet 2022;54(9):1376–89.

40. Villani A, Davidson S, Kanwar N, et al. The clinical utility of integrative genomics in childhood cancer extends beyond targetable mutations. Nat Cancer 2023;4(2):203–21.

41. Umeda M, Ma J, Huang BJ, et al. Integrated Genomic Analysis Identifies UBTF Tandem Duplications as a Recurrent Lesion in Pediatric Acute Myeloid Leukemia. Blood Cancer Discov 2022;3(3):194–207.

42. Schwartz JR, Ma J, Lamprecht T, et al. The genomic landscape of pediatric myelodysplastic syndromes. Nat Commun 2017;8(1):1557.

43. Xia J, Miller CA, Baty J, et al. Somatic mutations and clonal hematopoiesis in congenital neutropenia. Blood 2018;131(4):408–16.

44. Myers KC, Furutani E, Weller E, et al. Clinical features and outcomes of patients with Shwachman-Diamond syndrome and myelodysplastic syndrome or acute myeloid leukaemia: a multicentre, retrospective, cohort study. Lancet Haematol 2020;7(3):e238–46.

45. Welch JS, Petti AA, Miller CA, et al. TP53 and decitabine in acute myeloid leukemia and myelodysplastic syndromes. N Engl J Med 2016;375(21):2023–36.

46. Welch JS, Westervelt P, Ding L, et al. Use of whole-genome sequencing to diagnose a cryptic fusion oncogene. JAMA 2011;305(15):1577–84.

47. Stubbins RJ, Korotev S, Godley LA. Germline CHEK2 and ATM Variants in Myeloid and Other Hematopoietic Malignancies. Curr Hematol Malig Rep 2022;17(4):94–104.

48. Janiszewska H, Bąk A, Skonieczka K, et al. Constitutional mutations of the CHEK2 gene are a risk factor for MDS, but not for de novo AML. Leuk Res 2018;70:74–8.

49. Stubbins RJ, Arnovitz S, Vagher J, et al. Predisposition to hematopoietic malignancies by deleterious germline CHEK2 variants. Leukemia [Internet] 2025 [cited 2025 May 13];Available from: 10.1038/s41375-025-02635-1

50. Guijarro F, López-Guerra M, Morata J, et al. Germ line variants in patients with acute myeloid leukemia without a suspicion of hereditary hematologic malignancy syndrome. Blood Adv 2023;7(19):5799–811.

51. Slavin TP, Tsang KWK, Longmate J, et al. Effect of germline ATM mutations on clonal hematopoiesis. J Clin Oncol 2019;37(15_suppl):1509–1509.

52. Ito K, Hirao A, Arai F, et al. Regulation of oxidative stress by ATM is required for self-renewal of haematopoietic stem cells. Nature 2004;431(7011):997–1002.

53. Stubbins RJ, Asom AS, Wang P, Lager AM, Gary A, Godley LA. Germline loss of function BRCA1 and BRCA2 mutations and risk of de novo hematopoietic malignancies. Haematologica [Internet] 2023;Available from: 10.3324/haematol.2022.281580

54. Shah PD, Nathanson KL. BRCA1/2 mutations and de novo hematologic malignancies: true, true and not clearly related. Haematologica 2024;109(1):21–2.

55. Tung NM, Boughey JC, Pierce LJ, et al. Management of hereditary breast cancer: American Society of Clinical Oncology, American Society for Radiation Oncology, and Society of Surgical Oncology guideline. J Clin Oncol 2020;38(18):2080–106.

56. Camacho-Arias M, Larraz J, Martín-López L, et al. Case of refractory classic Hodgkin lymphoma with a germline pathogenic monoallelic variant in the TNFRSF13B gene. Pediatr Blood Cancer 2025;72(3):e31505.

57. Jasek M, Bojarska-Junak A, Sobczyński M, et al. Association of common variants of TNFSF13 and TNFRSF13B genes with CLL risk and clinical picture, as well as expression of their products-APRIL and TACI molecules. Cancers (Basel) 2020;12(10):2873.

58. Went M, Duran-Lozano L, Halldorsson GH, et al. Deciphering the genetics and mechanisms of predisposition to multiple myeloma. Nat Commun 2024;15(1):6644.

59. Verhoeven D, Stoppelenburg AJ, Meyer-Wentrup F, Boes M. Increased risk of hematologic malignancies in primary immunodeficiency disorders: opportunities for immunotherapy. Clin Immunol 2018;190:22–31.

60. Cumbo C, Orsini P, Tarantini F, et al. TNFRSF13B gene mutation in familial acute myeloid leukemia: A new piece in the complex scenario of hereditary predisposition? Hematol Oncol 2023;41(5):942–6.

61. Salzer U, Chapel HM, Webster ADB, et al. Mutations in TNFRSF13B encoding TACI are associated with common variable immunodeficiency in humans. Nat Genet 2005;37(8):820–8.

62. Lee JJ, Rauter I, Garibyan L, et al. The murine equivalent of the A181E TACI mutation associated with common variable immunodeficiency severely impairs B-cell function. Blood 2009;114(11):2254–62.

63. Romberg N, Virdee M, Chamberlain N, et al. TNF receptor superfamily member 13b (TNFRSF13B) hemizygosity reveals transmembrane activator and CAML interactor haploinsufficiency at later stages of B-cell development. J Allergy Clin Immunol 2015;136(5):1315–25.

64. Chapellier M, Peña-Martínez P, Ramakrishnan R, et al. Arrayed molecular barcoding identifies TNFSF13 as a positive regulator of acute myeloid leukemia-initiating cells. Haematologica 2019;104(10):2006–16.

65. Danishevich A, Bilyalov A, Nikolaev S, et al. CDKN2A gene mutations: Implications for hereditary cancer syndromes. Biomedicines 2023;11(12):3343.

66. Teierle SM, Huang Y, Kittai AS, et al. Characteristics and outcomes of patients with CLL and CDKN2A/B deletion by fluorescence in situ hybridization. Blood Adv 2023;7(23):7239–42.

67. Zhang W, Kuang P, Liu T. Prognostic significance of CDKN2A/B deletions in acute lymphoblastic leukaemia: a meta-analysis. Ann Med 2019;51(1):28–40.

68. Bertolotto C, Lesueur F, Giuliano S, et al. A SUMOylation-defective MITF germline mutation predisposes to melanoma and renal carcinoma. Nature 2011;480(7375):94–8.

69. Bonet C, Luciani F, Ottavi J-F, et al. Deciphering the role of oncogenic MITFE318K in senescence delay and melanoma progression. J Natl Cancer Inst [Internet] 2017 [cited 2025 Feb 14];109(8). Available from: https://pubmed.ncbi.nlm.nih.gov/28376192/

70. Morgan RD, Burghel GJ, Flaum N, et al. Extended panel testing in ovarian cancer reveals BRIP1 as the third most important predisposition gene. Genet Med 2024;26(10):101230.

71. Greaves M. A causal mechanism for childhood acute lymphoblastic leukaemia. Nat Rev Cancer 2018;18(8):471–84.

