## Supplement for "Germline Variant Burden Warrants Universal Genetic Testing in Pediatric Myeloid Leukemia"

**Supplementary Methods:** 2
Whole genome sequencing (WGS) 2
RNA sequencing 2
Cytogenetic and structural variants 2
Statistical Methods 2
**Supplementary Figures** 3
Supplementary Figure 1: Cohort Demographics 3
Supplementary Figure 2: Flowchart of programmatic filtering of variants to be hand-curated 4
Supplementary Figure 3: Demographics of germline variant carriers 5
Supplementary Figure 4: Oncoplot of germline and somatic variants 6
Supplementary Figure 5: Allelic bias in P/LP variants 7
Supplementary Figure 6: Number of variants in selected genes for 1000 Genomes Control subjects as compared to pediatric AML patients 8
**Supplementary Tables** 9
Supplementary Table 1: Cohort Demographics 9
Supplementary Table 2: Target genes associated with hematologic malignancies, cancer risk, immunodeficiencies, bone marrow, or blood diseases and genomic GRCh38 coordinates 10
Supplementary Table 3: P/LP variants with moderate evidence for association to hematopoietic malignancy risk 28
Supplementary Table 4: P/LP variants in genes with limited evidence for association to hematopoietic malignancy risk 30
Supplementary Table 5: P/LP variants with limited evidence for association to solid tumor risk 31
Supplementary Table 6: Heterozygous P/LP variants in genes with autosomal recessive hematopoietic malignancy risk inheritance 32
Supplementary Table 7: Heterozygous P/LP variants in genes with autosomal recessive solid tumor risk inheritance 33
Supplementary Table 8: Heterozygous P/LP variants in genes associated with other hematopoietic diseases or developmental disorders with autosomal recessive inheritance 35
Supplementary Table 9: P/LP variants in genes with a known somatic role in AML but no prior evidence for germline risk to malignancy 36
Supplementary Table 10: Patients with multiple P/LP variants 38
Supplementary Table 11: Additional information for P/LP variants 39
Supplementary Table 12: Additional somatic variants in genes with P/LP variants 48
Supplementary Table 13: Genes and mode of inheritance included in cohort meta-analysis 48
**Supplementary References:** 52

### Supplementary Methods:

#### Whole genome sequencing (WGS)

Whole-genome DNA sequencing was conducted as described previously[1]. Briefly, paired-end libraries were prepared using the Illumina library construction protocol and sequenced with 30x average coverage on Illumina HiSeq2500 instruments. Reads were mapped to GRCh37 (hg19) using bwa[2]; SNVs and indel variants were called with *Bambino*[3]. WGS for TARGET-21, previously aligned to GRCh38, was retrieved with BamSliceR[4].

#### RNA sequencing

Transcriptome data were generated by the British Columbia Genome Sciences Center (BCGSC; Vancouver, BC). Total RNA was ribo-depleted and reverse transcribed in a strand-specific mRNA library construction protocol, indexed, pooled, and sequenced on an Illumina HiSeq 2500 as previously published[5] to produce 75 base-pair, paired-end sequence reads. Sequencing data were aligned to human genome assembly GRCh38. Variants in regions of interest were identified using BamSliceR[4].

#### Cytogenetic and structural variants

Karyotypes were centrally reviewed at COG. Somatic structural variants for comparison between groups were called from WGS data using Manta with default parameters for somatic structural variant detection[6]. Suspected driver lesions were orthogonally validated as previously published.[7]

#### Statistical Methods

All statistical analyses were performed using R version 4.3.2. Figures were generated using ggplot21. Pathway analyses were performed using clusterProfiler[8]. Poisson goodness of fit tests were performed using vcd (visualizing categorical data)[9]. The oncoplot of variants was generated using maftools[10].

### Supplementary Figures

#### Supplementary Figure 1: Cohort Demographics.

A) Ethnic classifications. B) Scatter plot of the distribution of ages. C) Sex. D) Major somatic driver groups.

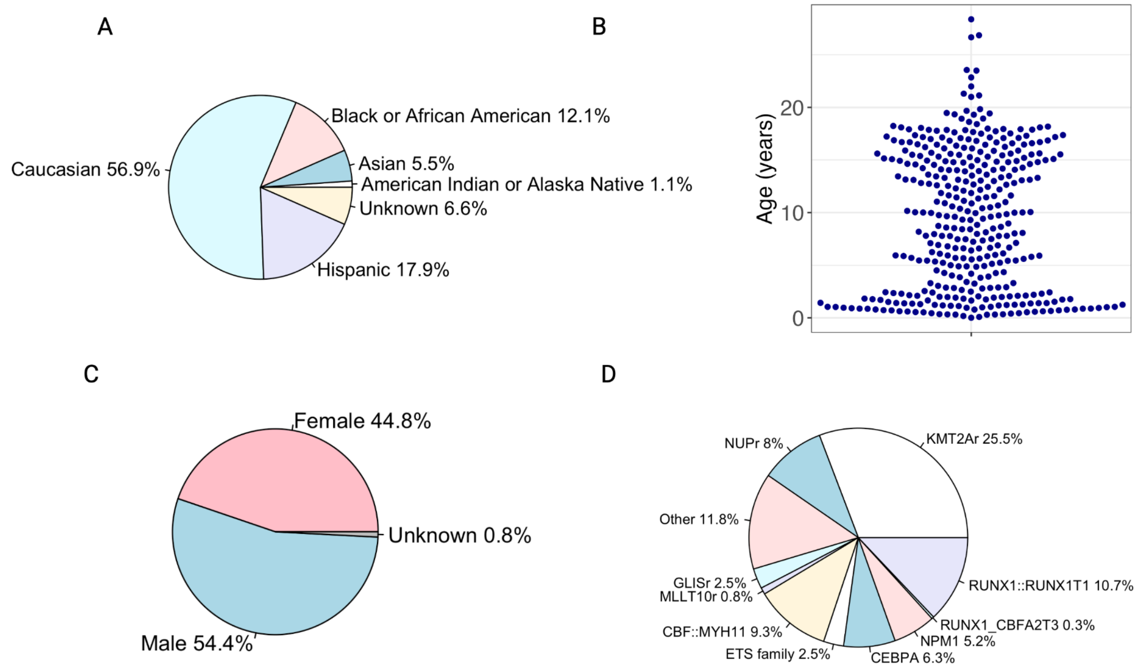

Supplementary Figure 2: Flowchart of programmatic filtering of variants to be hand-curated.

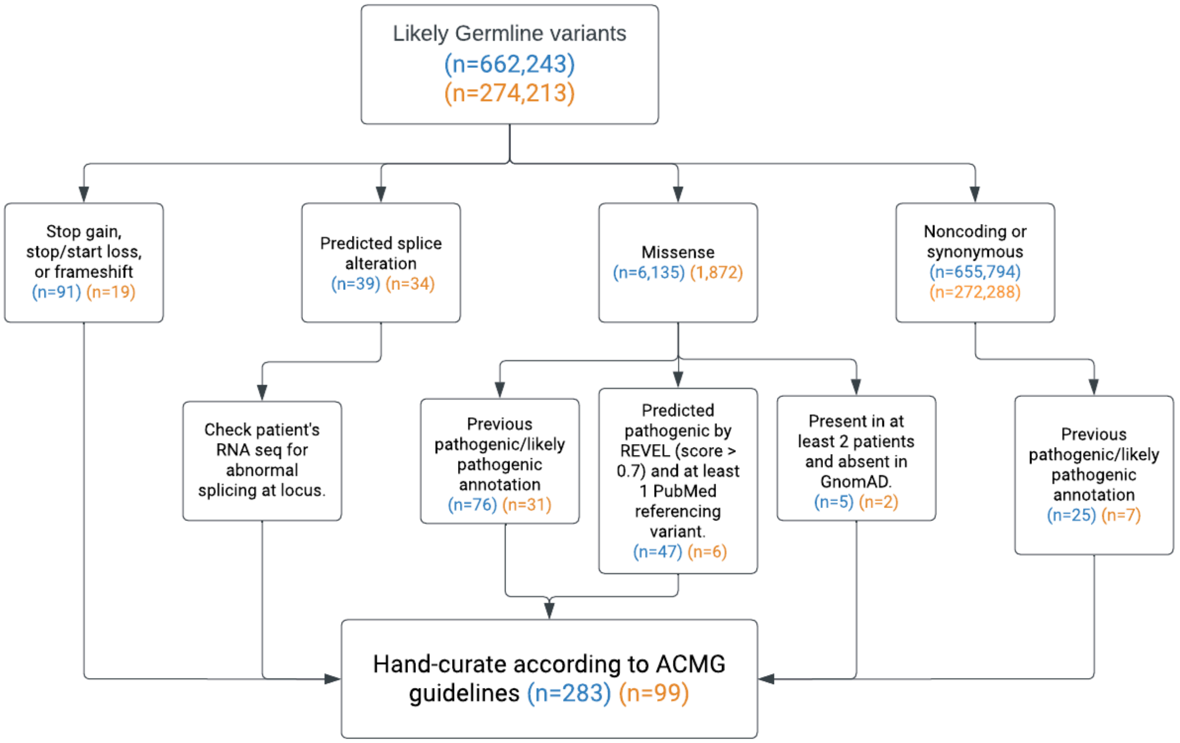


AAML1031 cohort (n=336 patients)
TARGET-21 cohort (n=29 patients)

Supplementary Figure 3: Demographics of germline variant carriers.

A-D) Comparison of patients carrying a germline variant versus patients without a germline variant. A) Distribution of ages, B) Sex, C) Ethnic classification, D) AML somatic driver group and E) Event-free survival.

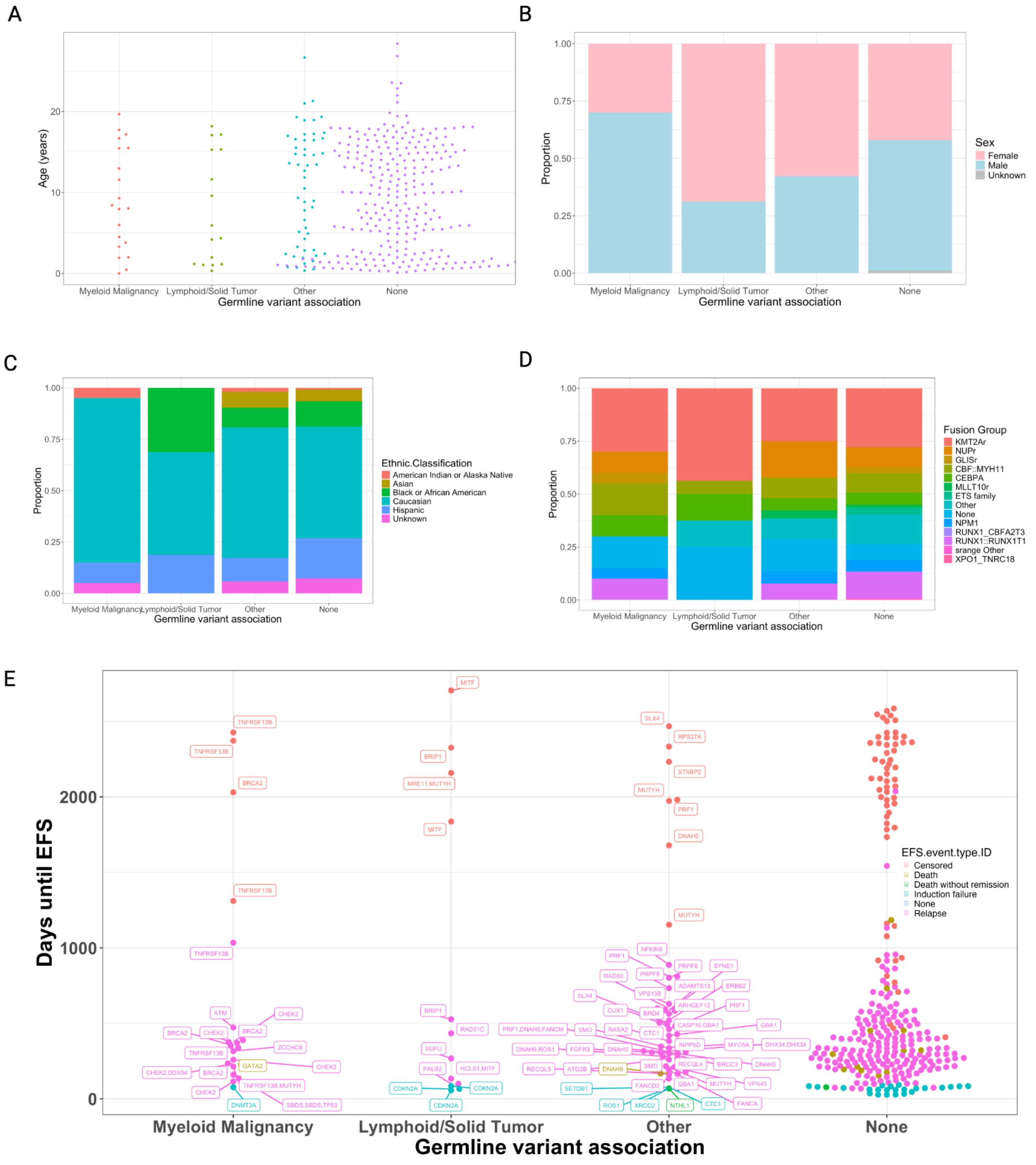

Supplementary Figure 4: Oncoplot of germline and somatic variants.

Columns represent patients. Green = germline variant calls, blue = somatic variants (covariates), red = somatic fusions (covariates), yellow = multiple variants.

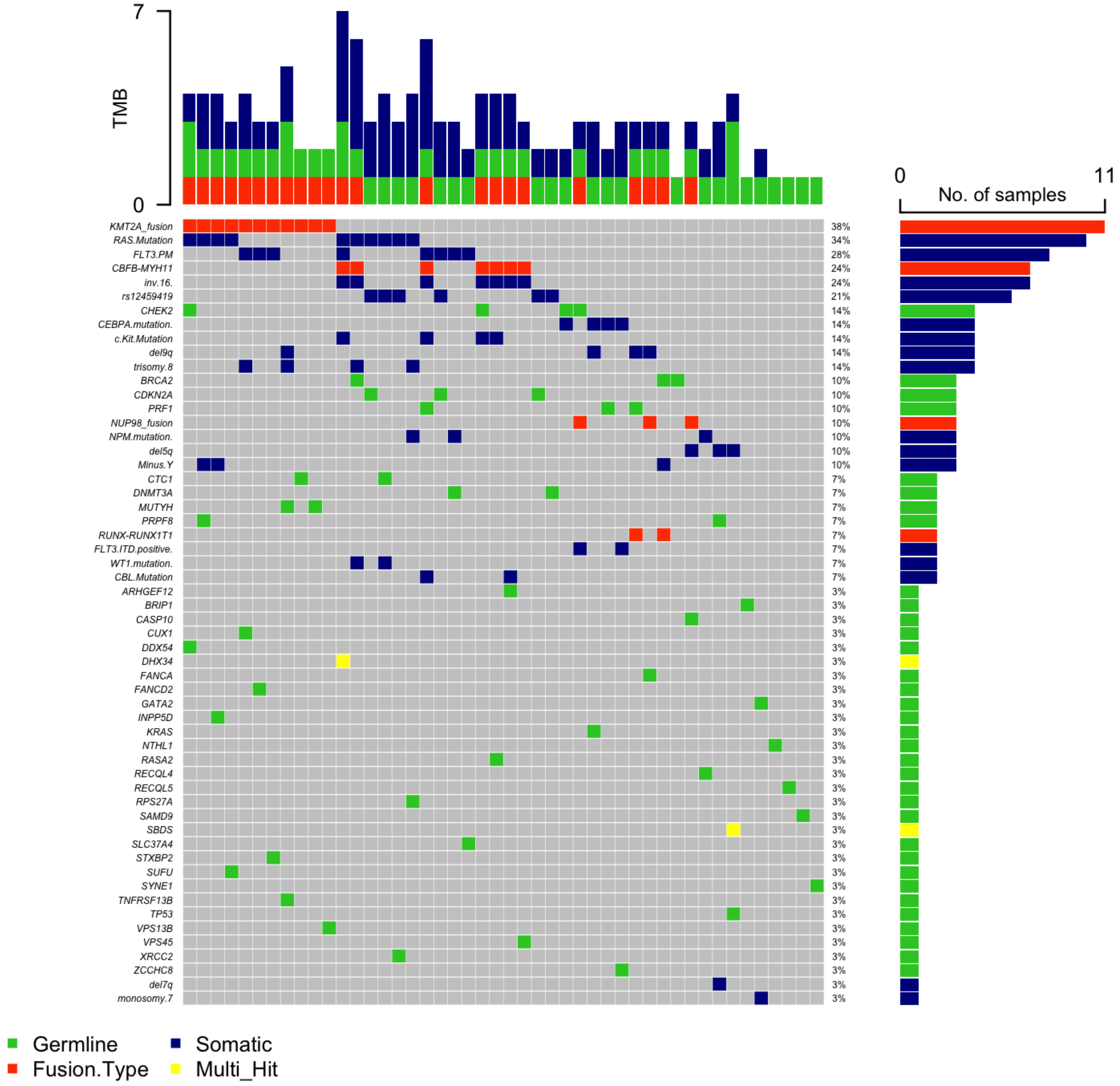

Supplementary Figure 5: Allelic bias in P/LP variants

A) P/LP variants and B) predicted nonsense-mediated mRNA decay (NMD) P/LP variants. VAF=variant allele frequency.

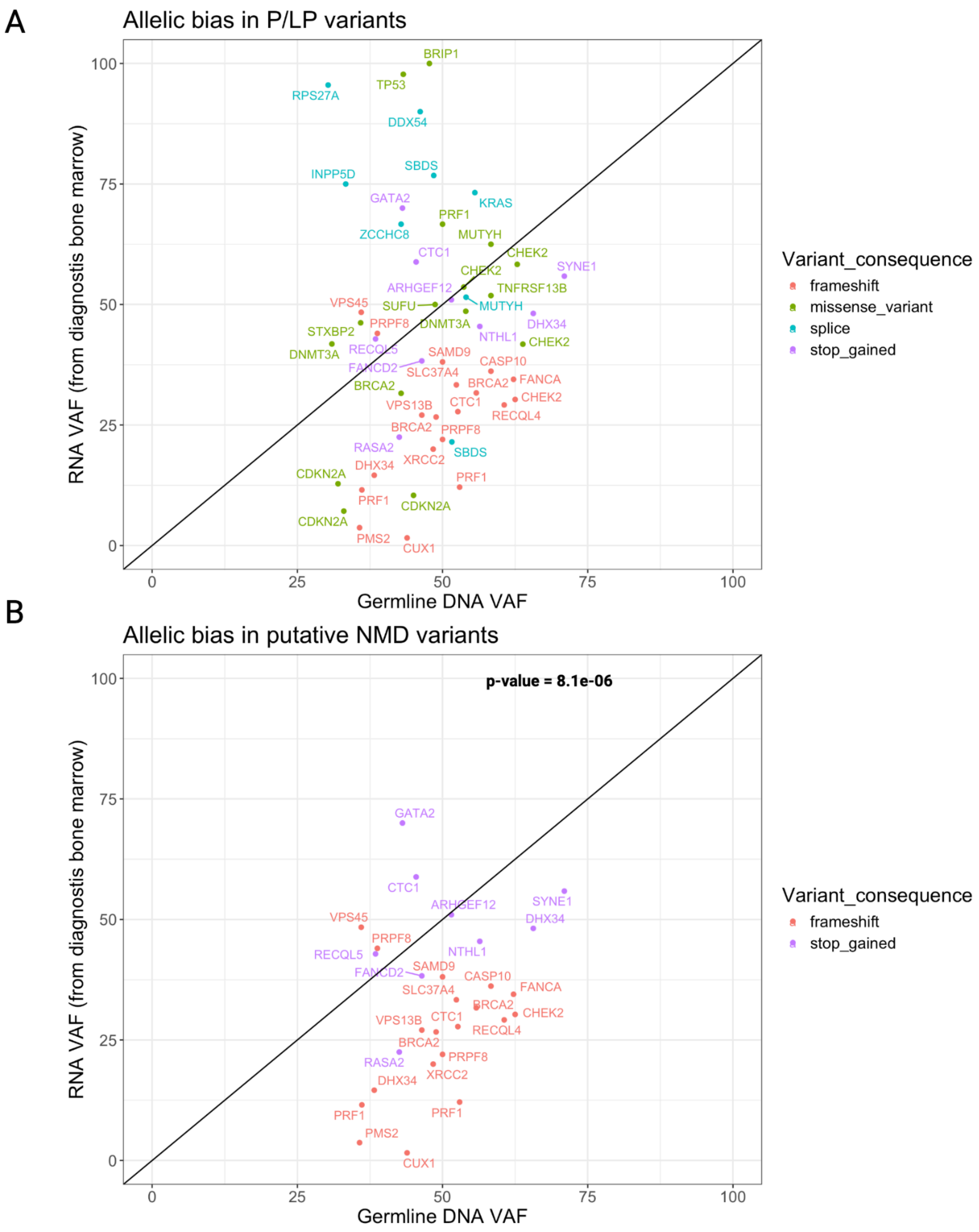

Supplementary Figure 6: Number of variants in selected genes for 1000 Genomes Control subjects as compared to pediatric AML patients

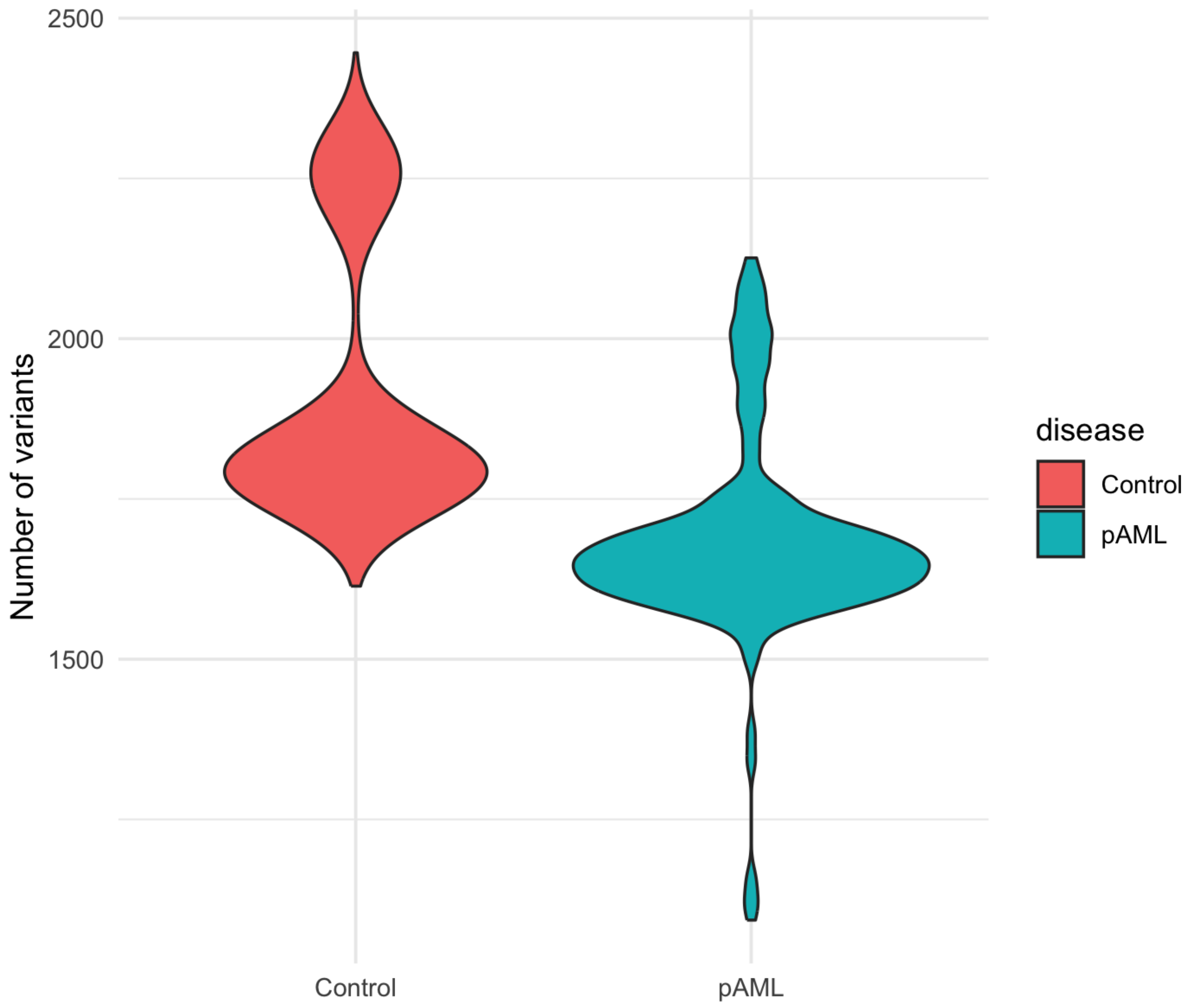

### Supplementary Tables

#### Supplementary Table 1: Cohort Demographics

| | AAML03P1 (N=4) | AAML0531 (N=25) | AAML1031 (N=335) | Overall (N=364) | p |
|---|---|---|---|---|---|
| **AgeGroup** | | | | | |
| AYA | 0 (0%) | 4 (16.0%) | 90 (26.9%) | 94 (25.8%) | 0.559 |
| Child | 3 (75.0%) | 16 (64.0%) | 152 (45.4%) | 171 (47.0%) | |
| Infant | 1 (25.0%) | 5 (20.0%) | 90 (26.9%) | 96 (26.4%) | |
| Missing | 0 (0%) | 0 (0%) | 3 (0.9%) | 3 (0.8%) | |
| **Sex** | | | | | |
| Female | 1 (25.0%) | 10 (40.0%) | 154 (46.0%) | 165 (45.3%) | 0.799 |
| Male | 3 (75.0%) | 15 (60.0%) | 181 (54.0%) | 199 (54.7%) | |
| **Fusion Group** | | | | | |
| CBF::MYH11 | 0 (0%) | 0 (0%) | 34 (10.1%) | 34 (9.3%) | <0.001 |
| CEBPA | 0 (0%) | 0 (0%) | 23 (6.9%) | 23 (6.3%) | |
| ETS family | 0 (0%) | 0 (0%) | 9 (2.7%) | 9 (2.5%) | |
| GLISr | 0 (0%) | 0 (0%) | 9 (2.7%) | 9 (2.5%) | |
| KMT2Ar | 0 (0%) | 2 (8.0%) | 101 (30.1%) | 103 (28.3%) | |
| MLLT10r | 1 (25.0%) | 2 (8.0%) | 3 (0.9%) | 6 (1.6%) | |
| NPM1 | 0 (0%) | 0 (0%) | 19 (5.7%) | 19 (5.2%) | |
| NUP98 NSD1/HOX | 2 (50.0%) | 5 (20.0%) | 16 (4.8%) | 23 (6.3%) | |
| NUP98 reader | 1 (25.0%) | 0 (0%) | 6 (1.8%) | 7 (1.9%) | |
| RUNX1::RUNX1T1 | 0 (0%) | 1 (4.0%) | 39 (11.6%) | 40 (11.0%) | |
| Other | 0 (0%) | 2 (8.0%) | 53 (15.8%) | 55 (15.1%) | |
| None | 0 (0%) | 13 (52.0%) | 23 (6.9%) | 36 (9.9%) | |
| **Ethnic Classification** | | | | | |
| Black or African American | 1 (25.0%) | 3 (12.0%) | 40 (11.9%) | 44 (12.1%) | 1 |
| Caucasian | 3 (75.0%) | 15 (60.0%) | 189 (56.4%) | 207 (56.9%) | |
| Asian | 0 (0%) | 2 (8.0%) | 18 (5.4%) | 20 (5.5%) | |
| Hispanic | 0 (0%) | 4 (16.0%) | 61 (18.2%) | 65 (17.9%) | |
| Unknown | 0 (0%) | 1 (4.0%) | 23 (6.9%) | 24 (6.6%) | |
| American Indian or Alaska Native | 0 (0%) | 0 (0%) | 4 (1.2%) | 4 (1.1%) | |

Supplementary Table 2: Target genes associated with hematologic malignancies, cancer risk, immunodeficiencies, bone marrow, or blood diseases and genomic GRCh38 coordinates.

Coordinates included all coding regions and untranslated regions (promoter, introns, 5’ and 3’ untranslated regions).

| **SYMBOL** | **Coordinates (GRCh38)** |
|---|---|
| ABL1 | 9:130713043-130887675 |
| ABL2 | 1:179099330-179229684 |
| ACD | 16:67657512-67660810 |
| ACTB | 7:5526409-5563902 |
| ACTN1 | 14:68874128-68979440 |
| ADA | 20:44584896-44652252 |
| ADA2 | 22:17178790-17258235 |
| ADAMTS13 | 9:133414358-133459402 |
| ADD3 | 10:109996368-110135565 |
| AIP | 11:67468174-67491154 |
| AIRE | 21:44285838-44298648 |
| AK2 | 1:33007986-33080996 |
| AKT1 | 14:104769349-104795759 |
| ALAS2 | X:55009055-55030977 |
| ALK | 2:29192774-29921586 |
| ANKRD26 | 10:26973793-27100494 |
| AP3B1 | 5:78000522-78294762 |
| APC | 5:112707518-112846239 |
| ARHGEF12 | 11:120336413-120489937 |
| ARID1A | 1:26693236-26782104 |
| ARID1B | 6:156776020-157210779 |
| ARID2 | 12:45729706-45908040 |
| ASXL1 | 20:32358330-32439319 |
| ASXL2 | 2:25733753-25878487 |
| ATF7IP | 12:14365676-14502931 |
| ATG2B | 14:96279195-96363341 |
| ATM | 11:108222804-108369102 |

| | |
|---|---|
| ATR | 3:142449007-142578733 |
| ATRX | X:77504880-77786233 |
| AXIN2 | 17:65528563-65561648 |
| BAP1 | 3:52401008-52410008 |
| BARD1 | 2:214725646-214809683 |
| BCL10 | 1:85265776-85276632 |
| BCL11B | 14:99169287-99272197 |
| BCL2 | 18:63123346-63320128 |
| BCL6 | 3:187721377-187745725 |
| BCOR | X:40049815-40177329 |
| BCORL1 | X:129981107-130058071 |
| BIRC3 | 11:102317484-102339403 |
| BIRC6 | 2:32357023-32619571 |
| BLM | 15:90717346-90816166 |
| BLOC1S3 | 19:45178784-45216933 |
| BLOC1S6 | 15:45587214-45615945 |
| BMPR1A | 10:86755786-86932825 |
| BRAF | 7:140719327-140924929 |
| BRCA1 | 17:43044295-43170245 |
| BRCA2 | 13:32315086-32400268 |
| BRCC3 | X:155071420-155123077 |
| BRD4 | 19:15235519-15332545 |
| BRINP3 | 1:190097658-190478404 |
| BRIP1 | 17:61679139-61863559 |
| BTG1 | 12:92140278-92145846 |
| BTK | X:101349338-101390796 |
| BUB1B | 15:40161023-40221123 |
| CALR | 19:12938578-12944489 |
| CAMTA1 | 1:6785454-7769706 |
| CARD11 | 7:2906142-3044228 |
| CASP10 | 2:201182872-201229428 |
| CASP8 | 2:201233443-201361836 |

| | |
|---|---|
| CBFA2T3 | 16:88874858-88977207 |
| CBFB | 16:67028984-67101058 |
| CBL | 11:119206298-119313926 |
| CBLB | 3:105655461-105869552 |
| CBLC | 19:44777869-44800652 |
| CCND1 | 11:69641156-69654474 |
| CCND3 | 6:41934934-42050357 |
| CCR4 | 3:32951644-32956349 |
| CD27 | 12:6444955-6451718 |
| CD40LG | X:136648158-136660390 |
| CD70 | 19:6583183-6604103 |
| CD79A | 19:41877279-41881372 |
| CD79B | 17:63928738-63932336 |
| CDAN1 | 15:42723544-42737128 |
| CDC73 | 1:193121983-193254815 |
| CDH1 | 16:68737292-68835537 |
| CDH11 | 16:64943753-65126112 |
| CDIN1 | 15:36579626-36810248 |
| CDK4 | 12:57747727-57756013 |
| CDKN1A | 6:36676460-36687337 |
| CDKN1B | 12:12685498-12722369 |
| CDKN1C | 11:2883213-2885775 |
| CDKN2A | 9:21967752-21995301 |
| CDKN2C | 1:50960745-50974634 |
| CEBPA | 19:33299934-33302534 |
| CEP57 | 11:95789965-95832693 |
| CHD2 | 15:92886203-93027996 |
| CHD4 | 12:6570082-6614524 |
| CHEK2 | 22:28687743-28742422 |
| CLPB | 11:72285495-72434680 |
| CREBBP | 16:3725054-3880713 |
| CRLF2 | Y:1190490-1212723, X:1190490-1212723 |

| | |
|---|---|
| CSF1R | 5:150053291-150113372 |
| CSF2RA | Y:1268800-1310381, X:1268800-1310381 |
| CSF3R | 1:36466043-36483278 |
| CTC1 | 17:8224815-8248058 |
| CTCF | 16:67562467-67639177 |
| CTLA4 | 2:203853888-203873965 |
| CTSC | 11:88265069-88359684 |
| CUX1 | 7:101815904-102283958 |
| CXCR2 | 2:218125289-218137251 |
| CXCR4 | 2:136114349-136119177 |
| CYCS | 7:25118656-25125260 |
| CYLD | 16:50742050-50801935 |
| DAXX | 6:33318558-33323016 |
| DDB2 | 11:47214465-47239217 |
| DDX41 | 5:177511577-177516961 |
| DDX54 | 12:113157173-113185479 |
| DHX15 | 4:24517441-24584554 |
| DHX29 | 5:55256055-55307694 |
| DHX34 | 19:47349315-47382704 |
| DICER1 | 14:95086228-95158010 |
| DIS3 | 13:72752169-72782096 |
| DIS3L2 | 2:231961245-232344350 |
| DKC1 | X:154762742-154777689 |
| DNAH5 | 5:13690328-14011818 |
| DNAH9 | 17:11598470-11969748 |
| DNAJC21 | 5:34929559-34958964 |
| DNM2 | 19:10718055-10833488 |
| DNMT1 | 19:10133342-10231286 |
| DNMT3A | 2:25227855-25342590 |
| DOCK8 | 9:214854-465259 |
| DROSHA | 5:31400494-31532196 |
| DTNBP1 | 6:15522807-15663058 |

| | |
|---|---|
| DTX1 | 12:113056730-113098028 |
| EBF1 | 5:158695920-159099916 |
| EED | 11:86201212-86278813 |
| EFL1 | 15:82130206-82262773 |
| EGFR | 7:55019017-55211628 |
| ELANE | 19:851014-856247 |
| EP300 | 22:41092510-41180077 |
| EPCAM | 2:47345158-47387601 |
| EPOR | 19:11377207-11384342 |
| ERBB2 | 17:39687914-39730426 |
| ERBB4 | 2:211375717-212538841 |
| ERCC1 | 19:45407334-45478828 |
| ERCC2 | 19:45349837-45370918 |
| ERCC3 | 2:127257290-127294166 |
| ERCC4 | 16:13920138-13952348 |
| ERCC5 | 13:102845831-102875995 |
| ERCC6L2 | 9:95871264-96121154 |
| ERG | 21:38380027-38661780 |
| ETNK1 | 12:22625075-22690665 |
| ETV6 | 12:11649674-11895377 |
| EXO1 | 1:241847967-241895148 |
| EXT1 | 8:117794490-118111826 |
| EXT2 | 11:44095648-44251962 |
| EZH2 | 7:148807257-148884321 |
| FADD | 11:70203296-70207390 |
| FAM111B | 11:59107185-59127412 |
| FAM47A | X:34129752-34132314 |
| FANCA | 16:89726683-89816977 |
| FANCB | X:14690388-14873255 |
| FANCC | 9:95099054-95426796 |
| FANCD2 | 3:10026370-10101932 |
| FANCE | 6:35452338-35467104 |

| | |
|---|---|
| FANCF | 11:22622533-22625823 |
| FANCG | 9:35073835-35080004 |
| FANCI | 15:89243945-89317261 |
| FANCL | 2:58159243-58241410 |
| FANCM | 14:45135930-45200890 |
| FAS | 10:88953813-89029605 |
| FASLG | 1:172659103-172666876 |
| FAT1 | 4:186587794-186726722 |
| FAT4 | 4:125314918-125492932 |
| FBXO11 | 2:47789316-47906498 |
| FBXW7 | 4:152320544-152536092 |
| FGFR1 | 8:38400215-38468834 |
| FGFR2 | 10:121478332-121598458 |
| FGFR3 | 4:1793293-1808872 |
| FH | 1:241497511-241519799 |
| FHIT | 3:59747277-61251459 |
| FLCN | 17:17212212-17237188 |
| FLI1 | 11:128686535-128813267 |
| FLT3 | 13:28003274-28100592 |
| FOXO1 | 13:40555667-40666641 |
| FOXO3 | 6:108559835-108684774 |
| FOXO4 | X:71095851-71103532 |
| FOXP1 | 3:70954693-71583978 |
| FYB1 | 5:39105252-39274528 |
| FYN | 6:111660332-111873452 |
| G6PC3 | 17:44070620-44082151 |
| GALE | 1:23795599-23800781 |
| GATA1 | X:48786540-48794311 |
| GATA2 | 3:128479427-128493201 |
| GATA3 | 10:8045378-8075198 |
| GBA1 | 1:155234452-155244699 |
| GFI1 | 1:92473043-92486925 |

| | |
|---|---|
| GFI1B | 9:132944000-132991687 |
| GINS1 | 20:25391008-25452700 |
| GNA13 | 17:65009289-65056740 |
| GNAS | 20:58839718-58911192 |
| GNB1 | 1:1785285-1892292 |
| GP1BA | 17:4932277-4935023 |
| GP1BB | 22:19723539-19724771 |
| GP9 | 3:129060779-129062406 |
| GPC3 | X:133535745-133987100 |
| GREM1 | 15:32718004-32745106 |
| GRHL2 | 8:101492439-101669726 |
| GSKIP | 14:96363452-96387288 |
| H1-4 | 6:26156329-26157115 |
| H3C11 | 6:27871845-27872346 |
| HAX1 | 1:154272355-154275875 |
| HCLS1 | 3:121631399-121660927 |
| HLTF | 3:149030127-149086554 |
| HNF1A | 12:120978543-121002512 |
| HNRNPK | 9:83968083-83980631 |
| HOXA11 | 7:27181157-27185232 |
| HOXB13 | 17:48724763-48728750 |
| HPS1 | 10:98410939-98446963 |
| HPS3 | 3:149129638-149173732 |
| HPS4 | 22:26443107-26483931 |
| HPS5 | 11:18278668-18322198 |
| HPS6 | 10:102065349-102068036 |
| HRAS | 11:532242-537321 |
| HVCN1 | 12:110627841-110704950 |
| ID3 | 1:23557926-23559501 |
| IDH1 | 2:208236229-208266074 |
| IDH2 | 15:90083045-90102477 |
| IFNGR2 | 21:33403413-33479348 |

| | |
|---|---|
| IGF2R | 6:159969082-160113507 |
| IGLL5 | 22:22887780-22896111 |
| IKZF1 | 7:50304068-50405101 |
| IKZF3 | 17:39757718-39864312 |
| IL17RA | 22:17084954-17115693 |
| IL2RG | X:71107404-71112108 |
| IL7R | 5:35852695-35879603 |
| INPP5D | 2:233059967-233207903 |
| IRF4 | 6:391739-411443 |
| IRF8 | 16:85899116-85922606 |
| ITGA2B | 17:44372180-44389649 |
| ITGB3 | 17:47253827-47313743 |
| ITK | 5:157142933-157255185 |
| JAGN1 | 3:9890574-9894349 |
| JAK1 | 1:64833223-65067754 |
| JAK2 | 9:4984390-5129948 |
| JAK3 | 19:17824780-17848071 |
| KDM6A | X:44873188-45112779 |
| KDR | 4:55078481-55125595 |
| KIF23 | 15:69414246-69448427 |
| KIT | 4:54657267-54740783 |
| KITLG | 12:88492793-88580851 |
| KLF1 | 19:12884422-12887201 |
| KLF2 | 19:16324826-16328685 |
| KLF6 | 10:3775996-3785281 |
| KLHDC8B | 3:49171598-49176486 |
| KLHL6 | 3:183487551-183555706 |
| KMT2A | 11:118436456-118526832 |
| KMT2C | 7:152134922-152436644 |
| KMT2D | 12:49018975-49060794 |
| KRAS | 12:25205246-25250936 |
| LAMTOR2 | 1:156054782-156058506 |

| | |
|---|---|
| LAPTM5 | 1:30732469-30757774 |
| LCK | 1:32251244-32286165 |
| LEF1 | 4:108047545-108168956 |
| LIG4 | 13:108207439-108218368 |
| LLGL2 | 17:75525080-75575209 |
| LMO2 | 11:33858576-33892076 |
| LRIG3 | 12:58872149-58920504 |
| LRRC4 | 7:128027071-128032107 |
| LUC7L2 | 7:139340359-139423457 |
| LYST | 1:235661041-235883724 |
| LZTR1 | 22:20982269-20999032 |
| MAD2L2 | 1:11658918-11691811 |
| MAF | 16:79585843-79600737 |
| MAFB | 20:40685848-40689236 |
| MAGT1 | X:77825747-77899271 |
| MAML1 | 5:179732822-179777283 |
| MAP2K1 | 15:66386837-66491656 |
| MAP2K2 | 19:4090321-4124122 |
| MAP2K4 | 17:12020829-12143830 |
| MAP3K14 | 17:45263119-45317029 |
| MASTL | 10:27154824-27187953 |
| MAX | 14:65006174-65102695 |
| MBD4 | 3:129430947-129440179 |
| MECOM | 3:169083499-169663775 |
| MED12 | X:71118543-71144103 |
| MEF2B | 19:19145567-19192131 |
| MEN1 | 11:64803510-64811294 |
| MET | 7:116672196-116798377 |
| MFHAS1 | 8:8783354-8893630 |
| MGA | 15:41621134-41773081 |
| MITF | 3:69739456-69968336 |
| MLH1 | 3:36993226-37050896 |

| | |
|---|---|
| MLH3 | 14:75013769-75051532 |
| MLLT10 | 10:21524646-21743630 |
| MN1 | 22:27748277-27801756 |
| MPL | 1:43337818-43354466 |
| MRE11 | 11:94415570-94493885 |
| MRTFA | 22:40410281-40636719 |
| MSH2 | 2:47403067-47663146 |
| MSH6 | 2:47695530-47810063 |
| MUC1 | 1:155185824-155192916 |
| MUTYH | 1:45329163-45340893 |
| MYB | 6:135181308-135219173 |
| MYC | 8:127735434-127742951 |
| MYCN | 2:15940550-15947007 |
| MYD88 | 3:38138552-38143024 |
| MYH9 | 22:36281280-36388010 |
| MYO5A | 15:52307281-52529132 |
| MYSM1 | 1:58643440-58700090 |
| NAF1 | 4:163110073-163166890 |
| NAPRT | 8:143574785-143578649 |
| NBEAL2 | 3:46979666-47009704 |
| NBN | 8:89924515-90003228 |
| NCOA4 | 10:46005088-46030623 |
| NCOR1 | 17:16029065-16218185 |
| NDRG1 | 8:133237175-133302022 |
| NF1 | 17:31094927-31382116 |
| NF2 | 22:29603553-29698598 |
| NFKBIE | 6:44258166-44265788 |
| NHP2 | 5:178149463-178153894 |
| NIPBL | 5:36876769-37066413 |
| NKX2-1 | 14:36516392-36521149 |
| NLRP2 | 19:54953130-55001142 |
| NOP10 | 15:34339159-34343180 |

| | |
|---|---|
| NOTCH1 | 9:136494433-136546048 |
| NOTCH2 | 1:119911553-120100779 |
| NPAT | 11:108157215-108222638 |
| NPM1 | 5:171387116-171411810 |
| NR4A3 | 9:99821855-99866891 |
| NRAS | 1:114704469-114716771 |
| NSD1 | 5:177131830-177300213 |
| NSD2 | 4:1871393-1982207 |
| NSUN2 | 5:6599239-6633291 |
| NT5C2 | 10:103087185-103277605 |
| NTHL1 | 16:2039815-2047866 |
| NTRK1 | 1:156815636-156881850 |
| NTRK3 | 15:87859751-88256791 |
| NUP98 | 11:3671083-3797792 |
| NXF1 | 11:62792123-62806302 |
| PALB2 | 16:23603160-23641321 |
| PARN | 16:14435700-14632728 |
| PAX5 | 9:36833269-37034268 |
| PBRM1 | 3:52545352-52685917 |
| PCLO | 7:82754012-83162930 |
| PDGFRA | 4:54229280-54298245 |
| PDGFRB | 5:150113839-150155872 |
| PGM3 | 6:83147324-83193936 |
| PHF6 | X:134373288-134428791 |
| PHOX2B | 4:41744082-41748725 |
| PIGA | X:15319452-15335554 |
| PIK3CD | 1:9629889-9729114 |
| PIK3R1 | 5:68215740-68301821 |
| PIM1 | 6:37170152-37175428 |
| PLCG1 | 20:41136960-41196801 |
| PLCG2 | 16:81779279-81962685 |
| PML | 15:73994673-74047827 |

| | |
|---|---|
| PMS1 | 2:189784085-189877629 |
| PMS2 | 7:5970925-6009130 |
| POLD1 | 19:50384204-50418018 |
| POLE | 12:132623753-132687376 |
| POLH | 6:43576185-43620523 |
| POT1 | 7:124822386-124929983 |
| PPM1D | 17:60600193-60666280 |
| PRDM1 | 6:105993463-106109939 |
| PRDM9 | 5:23443586-23528093 |
| PRF1 | 10:70597348-70602759 |
| PRKACG | 9:69012504-69014113 |
| PRKAR1A | 17:68511780-68551319 |
| PRKCB | 16:23835983-24220611 |
| PRPF40B | 12:49568218-49644666 |
| PRPF8 | 17:1650629-1684867 |
| PRPS1 | X:107628428-107651993 |
| PSMB5 | 14:23016543-23035230 |
| PTCH1 | 9:95442980-95517057 |
| PTEN | 10:87862638-87971930 |
| PTPN11 | 12:112418351-112509918 |
| PTPN2 | 18:12785478-12929643 |
| PTPRD | 9:8314246-10613002 |
| PTPRJ | 11:47980425-48170839 |
| RAB27A | 15:55202966-55319113 |
| RAC2 | 22:37225270-37259594 |
| RAD21 | 8:116845934-116874776 |
| RAD50 | 5:132556019-132646349 |
| RAD51 | 15:40694774-40732340 |
| RAD51C | 17:58692573-58735611 |
| RAD51D | 17:35092221-35121522 |
| RAF1 | 3:12582101-12664201 |
| RAG1 | 11:36510372-36593156 |

| | |
|---|---|
| RASA2 | 3:141487027-141615344 |
| RB1 | 13:48303744-48599436 |
| RBBP6 | 16:24537693-24572863 |
| RBM8A | 1:145921556-145927678 |
| RECQL | 12:21468910-21501669 |
| RECQL4 | 8:144511288-144517845 |
| RECQL5 | 17:75626845-75667189 |
| RELN | 7:103471381-103989658 |
| REST | 4:56907876-56966808 |
| RFWD3 | 16:74621399-74666877 |
| RHBDF2 | 17:76470891-76501790 |
| RHOA | 3:49359139-49412998 |
| RIF1 | 2:151409883-151508013 |
| RIT1 | 1:155897808-155911404 |
| RMRP | 9:35657754-35658017 |
| RNF168 | 3:196468783-196503768 |
| ROS1 | 6:117287353-117425942 |
| RPL10 | X:154389955-154409168 |
| RPL11 | 1:23691742-23696835 |
| RPL15 | 3:23916591-23924374 |
| RPL18 | 19:48615328-48619184 |
| RPL22 | 1:6185020-6209389 |
| RPL23 | 17:38847860-38853764 |
| RPL26 | 17:8377516-8383213 |
| RPL27 | 17:42998273-43002959 |
| RPL31 | 2:101002229-101024032 |
| RPL35 | 9:124857880-124861981 |
| RPL35A | 3:197950190-197956610 |
| RPL36 | 19:5674947-5691875 |
| RPL5 | 1:92832013-92841924 |
| RPS10 | 6:34417454-34426069 |
| RPS15 | 19:1438358-1440495 |

| | |
|---|---|
| RPS17 | 15:82536750-82540459 |
| RPS19 | 19:41860255-41872925 |
| RPS20 | 8:56067254-56074510 |
| RPS24 | 10:78033760-78056813 |
| RPS26 | 12:56041351-56044697 |
| RPS27 | 1:153990762-153992155 |
| RPS27A | 2:55231903-55235853 |
| RPS28 | 19:8321158-8323340 |
| RPS29 | 14:49570984-49599164 |
| RPS7 | 2:3575260-3580920 |
| RRAS | 19:49635292-49640143 |
| RTEL1 | 20:63657810-63696253 |
| RUNX1 | 21:34787801-36004667 |
| SAMD9 | 7:93099513-93118023 |
| SAMD9L | 7:93130056-93148385 |
| SAMHD1 | 20:36890229-36951893 |
| SBDS | 7:66987680-66995693 |
| SBF2 | 11:9776776-10304877 |
| SDHA | 5:218303-257082 |
| SDHAF2 | 11:61430042-61446839 |
| SDHB | 1:17018664-17054151 |
| SDHC | 1:161314381-161363206 |
| SDHD | 11:112086824-112120016 |
| SEC23B | 20:18507520-18561415 |
| SETBP1 | 18:44680173-45068510 |
| SETD2 | 3:47016428-47164113 |
| SETDB1 | 1:150926263-150964744 |
| SF1 | 11:64764606-64778786 |
| SF3A1 | 22:30331988-30356919 |
| SF3B1 | 2:197388515-197435079 |
| SH2B3 | 12:111405923-111451623 |
| SH2D1A | X:124227868-124373197 |

| | |
|---|---|
| SHANK2 | 11:70467854-71252577 |
| SHOC2 | 10:110919367-111017307 |
| SLC37A4 | 11:119023751-119030906 |
| SLFN14 | 17:35543985-35560819 |
| SLX4 | 16:3581181-3611606 |
| SMAD4 | 18:51028528-51085045 |
| SMARCA2 | 9:1980290-2193624 |
| SMARCA4 | 19:10960932-11079426 |
| SMARCB1 | 22:23786931-23838009 |
| SMARCD2 | 17:63832081-63843065 |
| SMC1A | X:53374149-53422728 |
| SMC3 | 10:110567684-110606048 |
| SMO | 7:129188633-129213545 |
| SOCS1 | 16:11254417-11256204 |
| SOS1 | 2:38962206-39124345 |
| SOS2 | 14:50117130-50231578 |
| SP140 | 2:230203110-230313215 |
| SPEN | 1:15836095-15940456 |
| SPI1 | 11:47354860-47409369 |
| SPRED1 | 15:38252836-38357249 |
| SRC | 20:37344685-37406050 |
| SRP54 | 14:34981957-35029686 |
| SRP72 | 4:56467617-56503681 |
| SRSF2 | 17:76734115-76737333 |
| STAG2 | X:123960212-124422664 |
| STAT3 | 17:42313324-42388568 |
| STAT5B | 17:42199176-42288633 |
| STIM1 | 11:3854527-4093210 |
| STK11 | 19:1177558-1228431 |
| STN1 | 10:103856806-103918332 |
| STX11 | 6:144150487-144191939 |
| STXBP2 | 19:7636772-7647873 |

| | |
|---|---|
| SUFU | 10:102503972-102633535 |
| SUZ12 | 17:31937007-32001038 |
| SYK | 9:90801787-90898549 |
| SYNE1 | 6:152121687-152637801 |
| TAFAZZIN | X:154411524-154421726 |
| TAL1 | 1:47216290-47232225 |
| TBL1XR1 | 3:177019340-177228000 |
| TCF3 | 19:1609291-1652615 |
| TCF4 | 18:55222185-55664787 |
| TCF7 | 5:134114681-134151865 |
| TCF7L2 | 10:112950247-113167678 |
| TCIRG1 | 11:68039025-68050895 |
| TERC | 3:169764520-169765060 |
| TERF2IP | 16:75647773-75761872 |
| TERT | 5:1253147-1295068 |
| TET2 | 4:105145875-105279816 |
| TFE3 | X:49028726-49043410 |
| THPO | 3:184371935-184381968 |
| TINF2 | 14:24238286-24242663 |
| TLX1 | 10:101131300-101137789 |
| TLX3 | 5:171309248-171312139 |
| TMEM127 | 2:96248514-96266047 |
| TNFAIP3 | 6:137867214-137883314 |
| TNFRSF13B | 17:16929816-16972118 |
| TNFRSF14 | 1:2555639-2565382 |
| TOX | 8:58805412-59119147 |
| TP53 | 17:7661779-7687546 |
| TPP1 | 11:6612768-6619448 |
| TRAF3 | 14:102777449-102911500 |
| TRIM24 | 7:138460259-138589996 |
| TSC1 | 9:132891348-132946874 |
| TSC2 | 16:2047967-2089491 |

| | |
|---|---|
| TSR2 | X:54440404-54448032 |
| TUBB1 | 20:59019429-59026654 |
| TYK2 | 19:10350533-10380608 |
| U2AF1 | 21:43092956-43107570 |
| U2AF2 | 19:55654146-55674716 |
| UBA2 | 19:34428352-34471251 |
| UBE2T | 1:202331544-202341984 |
| UBR5 | 8:102252273-102412759 |
| UNC13D | 17:75827225-75844785 |
| UROS | 10:125784980-125823288 |
| USB1 | 16:57999546-58021618 |
| USP7 | 16:8892097-8975328 |
| USP9X | X:41085445-41236579 |
| VAV1 | 19:6772708-6857366 |
| VHL | 3:10141778-10153667 |
| VPS13B | 8:99013266-99877580 |
| VPS45 | 1:150067279-150145329 |
| VWF | 12:5948877-6124770 |
| WAC | 10:28532493-28623112 |
| WAS | X:48676596-48691431 |
| WDR1 | 4:10068089-10116972 |
| WIF1 | 12:65050626-65121305 |
| WIPF1 | 2:174559572-174682916 |
| WRAP53 | 17:7686071-7703502 |
| WRN | 8:31033788-31176138 |
| WT1 | 11:32387775-32435564 |
| XBP1 | 22:28794555-28800597 |
| XIAP | X:123859712-123913972 |
| XPA | 9:97674909-97697340 |
| XPC | 3:14145147-14178621 |
| XPO1 | 2:61476032-61538741 |
| XRCC2 | 7:152644776-152676193 |

| | |
|---|---|
| ZBTB16 | 11:114059041-114256765 |
| ZBTB7A | 19:4043303-4066899 |
| ZCCHC8 | 12:122471600-122500932 |
| ZEB2 | 2:144364364-144521057 |
| ZFP36L2 | 2:43222402-43226606 |
| ZNF217 | 20:53567071-53609907 |
| ZNF384 | 12:6666477-6689572 |
| ZRSR2 | X:15790156-15830694 |

Supplementary Table 3: P/LP variants with moderate evidence for association to hematopoietic malignancy risk.

| Gene | Variant Type | Variant cDNA | Variant protein | ACMG/AMP Pathogenicity | ACMG/AMP Codes Applied | Germline Source | Classified in ClinVar | Summary of literature for germline risk |
|---|---|---|---|---|---|---|---|---|
| *ERBB2* | Frameshift | c.30dup | p.(Leu11AlafsTer99) | LP | PM2 supporting, PVS1 | BMRS | No | *ERBB2* variant identified to segregate with disease in a family with MPN/Melanoma. Odds ratio of 3.5 for MPN compared to healthy controls[11]. *ERBB2* variants were found in 2 cases with familial cancer (multiple types)[12]. They were also found in breast cancer family[13]. However, all these variants reported are missense and some are reported to stabilize the protein and activate HER2 signaling. |
| *CASP10* | Frameshift | c.947-950del | p.(Trp316LeufsTer11) | LP | PVS1, PM2 supporting | BMRS | No | *CASP10* is associated with Autoimmune lymphoproliferative syndrome (ALPS) with autosomal dominant inheritance. ALPS is associated with cancer risk, most strongly with lymphoma[14,15]. |
| *DHX34* | Stopgain[+] | c.1621C>T | p.(Arg541Ter) | LP | PVS1, PM2 supporting | BMRS | No | *DHX34* is involved in NMD, splicing, and hematopoietic differentiation. It has been previously connected to 4 families with MDS/AML, and functional studies showed that the variant abrogates its activity in NMD[16]. Germline missense and frameshift VUS were also identified in 3.2% of patients with marrow hypocellularity[17]. Additional functional assays show that loss of DHX34 regulates splicing of pre-mRNAs and results in differentiation blockade of erythroid and myeloid lineages[18]. |
| | Frameshift[+] | c.2463_2469del | p.(Ser821ArgfsTer50) | LP | PVS1, PM2 supporting | BMRS | No | |
| *PRF1* | Frameshift | c.50del | p.(Leu17ArgfsTer34) | P | PVS1, PS3, PS4 | BMRS | Yes | Homozygous loss of *PRF1* leads to familial hemophagocytic lymphohystiocytosis in infants. There is evidence that partial loss of *PRF1* can lead to delayed-onset FHL and hematopoietic malignancies[19]. Another paper found higher incidence of *PRF1* variants than normal controls in patients with Anaplastic large cell lymphoma[20]. Two other case reports documented hemizygous *PRF1* variants in cases of FLH[21,22]. |
| | Frameshift | c.50del | p.(Leu17ArgfsTer34) | P | PVS1, PS3, PS4 | BMRS | Yes | |
| | Frameshift | c.50del | p.(Leu17ArgfsTer34) | P | PVS1, PS3, PS4 | BMRS | Yes | |
| | Missense | c.666C>A | p.(His222Gln) | P | PS4, PS3, PM1, PP3 | BMRS | Yes | |

| | | | | | | | | |
|---|---|---|---|---|---|---|---|---|
| *FANCA* | Frameshift | c.1615del | p.(Asp539ThrfsTer66) | LP | PVS1, PM2 supporting | BMRS | Yes | *FANCA* heterozygous variants are generally not thought to cause disease. A study of 396 Fanconi anemia relatives that were heterozygous carriers did not find increased overall cancer risks, although sample sizes were underpowered for evaluating individual genes[23]. However, one study found *FANCA* variants significantly enriched in aplastic anemia and AML patients[24]. Another study found heterozygous *FANCA* variants at a higher rate in AML than would be expected by chance[25]. Other studies indicate a moderate increase in risk to breast cancer [26–28] |
| *RASA2* | Stopgain | c.874C>T | p.(Gln292Ter) | LP | PVS1, PM2 supporting | BMRS | No | *RASA2* loss of function variants were seen in three patients with Noonan syndrome that otherwise lacked a variant in a Noonan syndrome associated gene. Functional studies showed that heterozygous knockdown of *RASA2* resulted in RAS-ERK pathway activation, further supporting a causative role[29]. *RASA2* is mutated somatically in 5% of melanomas and also present in other cancers[30]. In COSMIC, it is mutated somatically in 0.85% of hematopoietic and lymphoid malignancies, further supporting its role as a tumor suppressor gene. |

ACMG - American College of Medical Genetics and Genomics, AMP - Association of Molecular Pathology, P - pathogenic, LP - likely pathogenic, AR - autosomal recessive, AD - autosomal dominant. BMRS - Bone marrow remission sample, BMSC - Bone marrow stromal cells. +Variants in the same patient, but not confirmed in trans.

Supplementary Table 4: P/LP variants in genes with limited evidence for association to hematopoietic malignancy risk.

| Gene | Variant Type | Variant cDNA | Variant protein | ACMG/AMP Pathogenicity | ACMG/AMP Codes Applied | Germline Source | Classified in ClinVar | Summary of literature for germline risk |
|---|---|---|---|---|---|---|---|---|
| *DDX54* | Splice | c.2196-2A>C | Retention of intron 17/19 | LP | PM2 supporting, PVS1 | BMRS | No | *DDX54* contributes to genomic stability. GWAS has shown an association between *DDX54* and hematopoietic malignancy[31]. |
| *DNAH5* | Stopgain | c.328A>T | p.(Lys110Ter) | LP | PVS1, PM2 supporting | BMRS | No | *DNAH5* germline variants were identified in adult AML patients[32]. However, LOF variants are somewhat common in gnomAD (522 individuals). Primary cilia are microtubule-based organelles found on the surface of almost all human bone marrow cells. Primary cilia respond to developmental signaling pathways like Hedgehog (Hh) and Wnt/β-catenin. AML cells show reduced frequency of primary cilia with abnormal morphology[33]. |
| | Stopgain | c.11725C>T | p.(Arg3909Ter) | LP | PVS1, PP4 | BMRS | Yes | |
| | Frameshift | c.13194_13197del | p.(Asp4398GlufsTer16) | P | PVS1, PM3, PP4 | BMRS | Yes | |
| | Stopgain | c.9502C>T | p.(Arg3168Ter) | LP | PVS1, PM2 supporting | BMRS | Yes | |
| *DNAH9* | Frameshift | c.308del | p.(Phe103SerfsTer31) | LP | PVS1, PM3 supporting | BMRS | Yes | Similarly to *DNAH5, DNAH9* germline variants were identified in adult AML patients[32]. AML cells show reduced frequency of primary cilia with abnormal morphology[33]. |
| | Stopgain | c.9421C>T | p.(Gln3141Ter) | LP | PVS1, PM2 supporting | BMRS | No | |
| *ATG2B* | Splice | c.5579+1G>T | Partial loss of exon 38/42. | LP | PVS1 (RNA), PM2 supporting | BMRS | No | *ATG2B* has been documented to contribute to familial MPNs with a germline duplication. Frameshift mutations in *ATG2B* are common in gastric cancers with high microsatellite instability[34]. *ATG2B* variants are predictive of survival in colorectal cancer[35]. *ATG2B* regulates cancer stemness in TNBC[36]. |

ACMG - American College of Medical Genetics and Genomics, AMP - Association of Molecular Pathology, P - pathogenic, LP - likely pathogenic, AR - autosomal recessive, AD - autosomal dominant. BMRS - Bone marrow remission sample, BMSC - Bone marrow stromal cells.

Supplementary Table 5: P/LP variants with limited evidence for association to solid tumor risk.

| Gene | Variant Type | Variant cDNA | Variant protein | ACMG/AMP Pathogenicity | ACMG/AMP Codes Applied | Germline Source | Classified in ClinVar | Summary of literature for germline risk |
|---|---|---|---|---|---|---|---|---|
| *SMO* | Missense | c.1921C>G | p.(Pro641Ala) | LP | PS3, PP1, PP3 | BMRS | Yes | Germline *SMO* P641A was found in a patient with non-small cell lung cancer, and his daughter had the variant and developed a skin basal cell carcinoma at a young age. Functional evidence showed activation of hedgehog pathway genes that was similar to overexpression of *SMO*[37]. |
| | Missense | c.1921C>G | p.(Pro641Ala) | LP | PS3, PP1, PP3 | BMRS | Yes | |
| *ROS1* | Frameshift | c.350del | p.(Leu117Tyrfs Ter11) | LP | PVS1, PM2 supporting | BMRS | No | *ROS1* germline variants were identified in two families with *BRCA1/2* negative breast cancer[38]. |
| | Stopgain | c.4190G>A | p.(Trp1397Ter) | LP | PVS1, PM2 supporting | BMRS | No | |
| *SLX4* | Frameshift | c.3895_3896del | p.(Arg1299GlyfsTer35) | LP | PVS1, PM2 supporting | BMSC | Yes | *SLX4* heterozygous variants are generally not thought to cause disease, although some publications show that *SLX4* heterozygous loss of function variants, although rare, have been found in cases of familial breast cancer [39,40] |
| | Stopgain | c.928C>T | p.(Arg310Ter) | LP | PVS1, PM2 supporting | BMRS | No | |
| *FANCM* | Stopgain | c.5791C>T | p.(Arg1931Ter) | LP | PVS1 strong, PS3-supporting, PM1 | BMRS | Yes | A large-scale analysis of >2000 familial BC without *BRCA1/2* found an increased OR for *FANCM* (OR, 2.44; 95% CI, 1.08-5.59; P = .02)[41]. This variant Arg1931Ter associates with ER-negative breast cancer risk (OR = 1.96; P = 0.006). The variant affected cell survival and chromosomal stability[42]. |
| *RECQL5* | Stopgain | c.2791A>T | p.(Lys931Ter) | LP | PVS1 (RNA), PM2 supporting | BMRS | No | *RECQL5* is involved in homologous recombination. It has been proposed to be involved in breast cancer susceptibility and was found in familial cases without *BRCA1/2*[43]. Another study found an association with *RECQL5* polymorphisms in a Chinese population of breast cancer patients[44]. |

ACMG - American College of Medical Genetics and Genomics, AMP - Association of Molecular Pathology, P - pathogenic, LP - likely pathogenic, AR - autosomal recessive, AD - autosomal dominant. BMRS - Bone marrow remission sample, BMSC - Bone marrow stromal cells.

Supplementary Table 6: Heterozygous P/LP variants in genes with autosomal recessive hematopoietic malignancy risk inheritance.

| Gene | Variant Type | Variant cDNA | Variant protein | ACMG/AMP Pathogenicity | ACMG/AMP Codes Applied | Germline Source | Classified in ClinVar | Summary of literature for germline risk |
|---|---|---|---|---|---|---|---|---|
| *CTC1* | Frameshift | c.1360del | p.(Glu454SerfsTer9) | LP | PVS1, PP4 | BMSC | Yes | Heterozygous *CTC1* variants may contribute to the onset of acquired BMF and clonal outgrowth of PNH clones. Heterozygous *CTC1 is* enriched in AA/PNH cases and associated with shortened telomeres (mean 7 as compared to mean 15 in age-matched controls). However, AA/PNH patients without telomere variants also had shortened telomeres[45]. |
| | Stopgain | c.277C>T | p.(Gln93Ter) | LP | PVS1, PM2 supporting | BMRS | Yes | |
| *FANCD2* | Stopgain | c.982C>T | p.(Arg328Ter) | LP | PVS1, PM2 supporting | BMRS | Yes | There is no published evidence of association of heterozygous *FANCD2* to familial cancer. |
| *RAD50* | Stopgain | c.832C>T | p.(Arg278Ter) | LP | PVS1, PP4 | BMRS | Yes | One study found *RAD50* variants are not associated with increased breast cancer risk, but with worse survival[46]. A large meta-analysis showed no increased risk of breast cancer or other cancers for *RAD50*[47]. |
| *RPS27A* | Splice | c.-17-98T>A | Retention of intron 1 (upstream of start) | LP | PVS1 (RNA), PM2 supporting | BMRS | No | There is no published evidence of heterozygous *RPS27A* variants in familial cancer. |
| *STXBP2* | Missense | c.1654G>A | p.(Gly552Ser) | LP | PS3, PM3, PP3 | BMRS | Yes | There is no published evidence of heterozygous *STXBP2* variants in familial cancer. |
| *VPS45* | Frameshift | c.695_696del | p.(Tyr232SerfsTer16) | LP | PVS1, PM2 supporting | BMRS | Yes | There is no published evidence of heterozygous *VPS45* variants in familial cancer. |
| *XRCC2* | Frameshift | c.96del | p.(Phe32LeufsTer30) | LP | PVS1, PM2 supporting | BMSC | Yes | *XRCC2* c.96delT is a founder variant in Poland. One study did not find an association to breast cancer[48]. |

ACMG - American College of Medical Genetics and Genomics, AMP - Association of Molecular Pathology, P - pathogenic, LP - likely pathogenic, AR - autosomal recessive, AD - autosomal dominant. BMRS - Bone marrow remission sample, BMSC - Bone marrow stromal cells.

Supplementary Table 7: Heterozygous P/LP variants in genes with autosomal recessive solid tumor risk inheritance.

| Gene | Variant Type | Variant cDNA | Variant protein | ACMG/AMP Pathogenicity | ACMG/AMP Codes Applied | Germline Source | Classified in ClinVar | Summary of literature for germline risk |
|---|---|---|---|---|---|---|---|---|
| *MUTYH* | Missense | c.1187G>A | p.(Gly396Asp) | P | PS3, PS4, PP3 | BMRS | Yes | Heterozygous variants in *MUTYH* are associated with a slightly increased risk of colorectal cancer[49]. Another large study found the following hazard ratios and 95% CIs for heterozygous *MUTYH* carriers: gastric cancer 9.3 (6.7-13); hepatobiliary cancer 4.5 (2.7-7.5); endometrial cancer 2.1 (1.1-3.9) and breast cancer 1.4 (1.0-2.0)[50]. Other studies also found elevated risk of liver and gastric cancers and a slightly increased risk of breast cancer for heterozygous *MUTYH* [51,52]. Another study confirmed a higher frequency of heterozygous *MUTYH* variants in individuals with cancer and attributed this to loss of heterozygosity of the functional allele[53]. |
| | Missense | c.1187G>A | p.(Gly396Asp) | P | PS3, PS4, PP3 | BMRS | Yes | |
| | Missense | c.1187G>A | p.(Gly396Asp) | P | PS3, PS4, PP3 | BMRS | Yes | |
| | Missense | c.1187G>A | p.(Gly396Asp) | P | PS3, PS4, PP3 | BMRS | Yes | |
| | Missense | c.536A>G | p.(Tyr179Cys) | P | PS3, PS4, PP3 | BMRS | Yes | |
| NTHL1 | Stopgain | c.835C>T | p.(Gln279Ter) | P | PVS1 strong, PM3 supporting, PS3 | BMRS | Yes | There is insufficient evidence of *NTHL1* heterozygous carriers having an increased risk of cancer, although one study found a low increase in breast cancer risk[54]. |

| | | | | | | | | |
|---|---|---|---|---|---|---|---|---|
| *RECQL4* | Frameshift | c.3072del | p.(Val1026Cy sfsTer18) | P | PVS1, PM2 supporting, PM3 supporting | BMRS | Yes | Homozygous variants in *RECQL4* cause Rothmund-Thomson syndrome, associated with increased risk of childhood cancer. Heterozygous loss of function variants in RECQL4 were enriched in pediatric osteosarcoma patients (odds ratio=7.1, 95% CI, 2.9-17)[55]. However, a study of 123 heterozygous *RECQL4* carriers did not show an increased incidence in cancer[56]. |
| XPC | Stopgain | c.463C>T | p.(Arg155Ter) | LP | PVS1, PM2 supporting | BMRS | Yes | *XPC* is known to confer cancer risk in homozygous form. One study showed that *XPC* polymorphism carriers have a slight increase in risk for breast, bladder, head and neck, and lung cancer[57]. A study of lung adenocarcinoma in mice showed a dose-dependent effect, where mice heterozygous for *XPC* developed an intermediate number of tumors when exposed to cigarette smoke[58]. *XPC* polymorphisms associated with survival in AML patients[59]. |

ACMG - American College of Medical Genetics and Genomics, AMP - Association of Molecular Pathology, P - pathogenic, LP - likely pathogenic, AR - autosomal recessive, AD - autosomal dominant. BMRS - Bone marrow remission sample, BMSC - Bone marrow stromal cells.

Supplementary Table 8: Heterozygous P/LP variants in genes associated with other hematopoietic diseases or developmental disorders with autosomal recessive inheritance.

| Gene | Variant Type | Variant cDNA | Variant protein | ACMG/AMP Pathogenicity | ACMG/AMP Codes Applied | Germline Source | Classified in ClinVar | Summary of literature for germline risk |
|---|---|---|---|---|---|---|---|---|
| *ADAMTS13* | Missense | c.559G>C | p.(Asp187His) | LP | PS3 moderate, PM1, PM3, PP3 | BMRS | Yes | Homozygous or compound heterozygous variants are associated with Thrombotic thrombocytopenic purpura, a bleeding disorder. Carriers of *ADAMTS13* are generally considered unaffected. However, one study did find decreased levels of *ADAMTS13* expression in ALL patients, and lower expression associated with patients with infections and high-risk status[60]. Low *ADAMTS13* levels were also found in AML patients and correlated with worse BMT outcomes[61]. |
| *GBA1* | Missense | c.1483G>C | p.(Ala495Pro) | P | PS3, PS4, PM3 | BMRS | Yes | There is moderate evidence for homozygous *GBA1* variants in hematopoietic malignancy risk[62], but heterozygous *GBA1* variants are not documented to increase cancer risk. However, one study found decreased *GBA1* expression in liver cancer patients, which associated with vascular invasion and advanced stage disease due to activation of the WNT signaling pathway[63]. |
| | Missense | c.882T>G | p.(His294Gln) | LP | PM3 Strong, PS4 Moderate | BMRS | Yes | |
| | Missense | c.1226A>G | p.(Asn409Ser) | LP | PS3 moderate, PS4, PM1, PM3, PP1, PP2 | BMRS | Yes | |
| *VPS13B* | Frameshift | c.5120_5124 del | p.(Lys1707Serfs Ter9) | LP | PVS1, PM2 supporting | BMRS | No | Homozygous *VPS13B* variants can cause Cohen syndrome, which involves neutropenia. There is no published evidence of heterozygous carriers being affected. |

ACMG - American College of Medical Genetics and Genomics, AMP - Association of Molecular Pathology, P - pathogenic, LP - likely pathogenic, AR - autosomal recessive, AD - autosomal dominant. BMRS - Bone marrow remission sample, BMSC - Bone marrow stromal cells.

Supplementary Table 9: P/LP variants in genes with a known somatic role in AML but no prior evidence for germline risk to malignancy.

| Gene | Variant Type | Variant cDNA | Variant protein | ACMG/AMP Pathogenicity | ACMG/AMP Codes Applied | Germline Source | Classified in ClinVar | Summary of literature for germline risk |
|---|---|---|---|---|---|---|---|---|
| *ARHGEF12* | Stopgain | c.76C>T | p.(Arg26Ter) | LP | PVS1, PM2 supporting | BMRS | No | *ARHGEF12* has a known somatic role in leukemia. Heterozygous loss of function variants are rare in GnomAD (20 individuals). |
| *CUX1* | Frameshift | c.1754del | p.(Phe585SerfsTer11) | LP | PVS1, PM2 supporting | BMRS | Yes | *CUX1* somatic variants are known to contribute to myeloid malignancy[64]. *CUX1* heterozygous germline variants cause mild to moderate intellectual disability/developmental delay of variable phenotype that resolves in adulthood[65]. Heterozygous loss of function variants are rare in GnomAD (11 individuals). |
| *PRPF8* | Frameshift | c.605_606del | p.(Pro202ArgfsTer7) | LP | PVS1, PM2 supporting | BMRS | No | *PRPF8* has a known somatic mechanism for AML and MDS. Its inheritance is AD for Retinitis pigmentosa, but average age of onset is 35. Heterozygous loss of function variants are rare in GnomAD (29 individuals). |
| | Frameshift | c.6991del | p.(Glu2331ArgfsTer28) | LP | PVS1 strong, PM2 supporting, PP1 | BMRS | Yes | |
| *BRCC3* | Splice | c.196-2del | Retention of intron 3/10. | LP | PVS1 (RNA), PM2 supporting, PP3 | BMRS | No | *BRCC3* is a component of the DNA repair pathway. Loss of function variants are rare in GnomAD (4 heterozyotes, 2 homozygotes). Somatic mutations have been reported in MDS and MPNs[66]. |
| *BRD4* | Splice | c.3782+2T>G | Loss of exon 18/20 | LP | PS3 (RNA), PM2 supporting, PP3 | BMSC | No | *BRD4* is well-known for somatic mutations in cancer. Heterozygous LOF variants cause pre- and postnatal growth defects [67]. Heterozygous loss of function variants are rare in GnomAD (2 individuals). |
| *FGFR3* | Splice | c.445+2T>G | Retention of intron 4/17 | LP | PVS1 (RNA), PM2 supporting | BMRS | No | *FGFR3* is mostly known for its somatic role. However it's been observed to accumulate in the male germline via clonal expansion in the testes and correlating with age[68]. Germline variants are associated with skeletal disorders. Heterozygous loss of function variants are rare in GnomAD (36 individuals). |
| *INPP5D* | Splice | c.349+2T>G | Retention of intron 3/26 | LP | PVS1 (RNA), PP3 | BMRS | No | *INPP5D* is known for its somatic role in myeloid malignancy. Mouse studies show complete knockout of *INPP5D* is early postnatal lethal due to overproliferation of myeloid cells[69]. The effects of heterozygous loss of function are unknown, although present in 50 GnomAD subjects. |

| | | | | | | | | |
|---|---|---|---|---|---|---|---|---|
| *MYO5A* | Stopgain | c.2416C>T | p.(Arg806Ter) | LP | PVS1, PM2 supporting | BMRS | No | *MYO5A* is associated with Griscelli syndrome with autosomal recessive inheritance. It has been reported as a fusion partner in pediatric low-grade glioma[70] and upregulation has been seen in melanoma and other cancer types[71]. Heterozygous loss of function variants are rare in GnomAD (36 individuals). |
| *NFKBIE* | Frameshift | c.137del | p.(Ile46ThrfsTer3) | LP | PVS1, PM2 supporting | BMRS | No | *NFKBIE* is commonly mutated somatically in B-cell malignancies and activates the NF-κB pathway[72]. Heterozygous loss of function variants are rare in GnomAD (1 individual). |
| *SETDB1* | Stopgain | c.1214C>A | p.(Ser405Ter) | LP | PVS1, PM2 supporting | BMSC | No | *SETDB1* has a known somatic role in cancer[73]. Heterozygous loss of function variants are rare in GnomAD (12 individuals). |
| *SYNE1* | Stopgain | c.23309T>A | p.(Leu7770Ter) | LP | PVS1, PM2 supporting | BMRS | No | *SYNE1* is commonly mutated somatically in cancer. *SYNE1* is associated with muscular dystrophy (AD) and spinocerebellar ataxia (AR). |

ACMG - American College of Medical Genetics and Genomics, AMP - Association of Molecular Pathology, P - pathogenic, LP - likely pathogenic, AR - autosomal recessive, AD - autosomal dominant. BMRS - Bone marrow remission sample, BMSC - Bone marrow stromal cells.

Supplementary Table 10: Patients with multiple P/LP variants.

| **USI** | **Genes with P/LP variant** |
|---|---|
| PATKWH | *BRD4, TRIM24* |
| PAUSBP | *CHEK2, DDX54* |
| PAUVWV | *MRE11A, MUTYH* |
| PAVCNG | *DNAH9, ROS1* |
| PAWPKR | *HCLS1, MITF* |
| PAWSJC | *DHX34, DHX34* |
| PAWUAE | *MUTYH, TNFRSF13B* |
| PAXELP | *DNAH5, FANCM, PRF1* |
| PAXJZX | *SBDS, SBDS, TP53* |

Supplementary Table 11: Additional information for P/LP variants.

| USI | Gene | Variant cDNA | Variant Protein | Variant Type | Transcript | ACMG Pathogenicity | ACMG Criteria | Tissue source | Fibroblast VAF | Remission VAF | Diagnosis VAF | Relapse VAF | RNA VAF | Sex | Age Group | Major Fusion or Somatic Driver | Fusion or Somatic Driver VAF in Remission WGS | MRD (%) at induction 1 | FAB | EFS event type ID |
|---|---|---|---|---|---|---|---|---|---|---|---|---|---|---|---|---|---|---|---|---|
| PAXLWH | ADAMTS13 | c.559G>C | p.(Asp187His) | Missense | NM_139025.5 | LP | PS3 moderate, PM1, PM3, PP3 | BMRS | NA | 0.61 | NA | 0.31 | 0.33 | M | Child | NUP98-NSD1 | 0 (FLT3-ITD) | 0 | M5 | Relapse |
| PAWZAU | ARHGEF12 | c.76C>T | p.(Arg26Ter) | Stopgain | NM_015313.3 | LP | PVS1, PM2 supporting | BMRS | NA | 0.52 | 0.38 | 0.61 | 0.51 | F | Child | CBFB-MYH11 | 0 | 0 | Unknown | Relapse |
| PAWVCG | ATG2B | c.5579+1G>T | Partial loss of exon 38/42. | Splice | NM_018036.7 | LP | PVS1 (RNA), PM2 supporting | BMRS | NA | 0.44 | 0.38 | 0.53 | 0* | M | Infant | KMT2A-MLLT3 | 0 | 0 | Unknown | Relapse |
| PAWWCW | ATM | c.8395_8404del | p.(Phe2799LysfsTer4) | Frameshift | NM_000051.4 | P | PVS1, PM2 supporting, PM3 | BMRS | NA | 0.37 | 0.21 | 0.41 | 0.32 | M | Infant | KMT2A_MLLT10 | 0 | 0 | M5 | Relapse |
| PAUWFM | BRCA2 | c.8377G>A | p.(Gly2793Arg) | Missense | NM-000059.4 | P | PS3, PS4, PM2-supporting, PP3 | BMRS | NA | 0.43 | 0.49 | NA | 0.32 | M | Infant | CBFA2T3-GLIS2 | +21 in tumor WGS and absent in remission WGS | 0.9 | M7 | Relapse |
| PAUXKI | BRCA2 | c.5410-5411del | p.(Val1804LysfsTer2) | Frameshift | NM-000059.4 | LP | PVS1, PM2-supporting | BMRS | NA | 0.49 | 0.5 | 0.4 | 0.27 | M | AYA | RUNX1-RUNX1T1 | 0 | 0 | Unknown | Relapse |
| PAVHWK | BRCA2 | c.5946del | p.(Ser1982ArgfsTer22) | Frameshift | NM-000059.4 | P | PVS1, PS3 | BMRS | NA | 0.56 | 0.41 | NA | 0.32 | F | Child | CBFB-MYH11 | 0 | 0 | Unknown | Censored |
| PAVKAU | BRCA2 | c.5614A>T | p.(Lys1872Ter) | Stopgain | NM-000059.4 | P | PVS1, PS4, PM2-supporting | BMRS | NA | 0.58 | 0.42 | 0.6 | NA | F | Infant | KMT2A-MLLT3 | 0 | 0 | Unknown | Relapse |
| PAVACZ | BRCC3 | c.196-2del | Retention of intron 3/10. | Splice | NM_024332.4 | LP | PVS1 (RNA), PM2 supporting, PP3 | BMRS | NA | 0.45 | 0.48 | NA | 0.4 | F | Child | KMT2A-MLLT3 | 0 | 0 | Unknown | Relapse |
| PATKWH | BRD4 | c.3782+2T>G | Loss of exon 18/20 | Splice | NM_001379291.1 | LP | PS3 (RNA), PM2 supporting, PP3 | BMSC | 0.3 | NA | NA | NA | 0* | M | AYA | None | NA | NA | M6 | Relapse |

| | | | | | | | | | | | | | | | | | | | | |
|---|---|---|---|---|---|---|---|---|---|---|---|---|---|---|---|---|---|---|---|---|
| PAUYSN | BRIP1 | c.2990-2993del | p.(Thr997ArgfsTer61) | Frameshift | NM-032043.3 | P | PVS1, PS4 | BMRS | NA | 0.45 | 0.43 | NA | 0* | M | Infant | None | Unknown | 0 | M5 | Censored |
| PAVDZK | BRIP1 | c.139C>G | p.(Pro47Ala) | Missense | NM-032043.3 | LP | PS3 moderate, PS4 moderate, PM1, PP3 | BMRS | NA | 0.48 | 0.91 | 1 | 1.00 | M | AYA | MED14-HOXA9 (promoter hijacking) | 0 | 0 | Unknown | Relapse |
| PAVEAF | CASP10 | c.947-950del | p.(Trp316LeufsTer11) | Frameshift | NM-032977.4 | LP | PVS1, PM2 supporting | BMRS | NA | 0.58 | 0.32 | NA | 0.36 | M | Child | NUP98-NSD1 | 0 | 7.4 | Unknown | Relapse |
| PARNAW | CDKN2A | c.194T>C | p.(Leu65Pro) | Missense | ENST00000304494.10 | LP | PS4, PP1 moderate, BP4 | BMSC | 0.45 | NA | NA | NA | 0.1 | F | Infant | None | NA | NA | M7 | Induction failure |
| PASFHK | CDKN2A | c.194T>C | p.(Leu65Pro) | Missense | ENST00000304494.10 | LP | PS4, PP1 moderate, BP4 | BMSC | 0.32 | NA | NA | NA | 0.13 | M | Child | None | NA | NA | M4 | Induction failure |
| PATAIJ | CDKN2A | c.194T>C | p.(Leu65Pro) | Missense | ENST00000304494.10 | LP | PS4, PP1 moderate, BP4 | BMSC | 0.33 | NA | NA | NA | 0.07 | F | Infant | KMT2A-MLLT10 | NA | NA | M5 | Induction failure |
| PAUSBP | CHEK2 | c.1100del | p.(Thr410MetfsTer15) | Frameshift | NM_007194.4 | P | PVS1, PS3 supporting, PS4 | BMRS | NA | 0.63 | NA | 0.43 | 0.30 | M | Child | KMT2A-AFDN | 17p deletion in tumor WGS and absent in remission WGS | 0 | M5 | Relapse |
| PAUVGD | CHEK2 | c.599T>C | p.(Ile157Thr) | Missense | NM_007194.4 | LP | PS3, PS4 moderate, PM1, BS1 | BMRS | NA | 0.64 | 0.6 | 0.58 | 0.42 | M | Child | NUP98-NSD1 | 0 | 12 | M0 | Relapse |
| PAWHML | CHEK2 | c.599T>C | p.(Ile157Thr) | Missense | NM_007194.4 | LP | PS3, PS4 moderate, PM1, BS1 | BMRS | NA | 0.54 | NA | 0.62 | 0.54 | M | AYA | CEBPA | 0 | 0 | M2 | Relapse |
| PAWHTL | CHEK2 | c.836T>C | p.(Ile157Thr) | Missense | NM_007194.4 | LP | PS3 supporting, PS4, PM1 | BMRS | NA | 0.5 | 0.48 | 0.44 | NA | M | Child | CBFB-MYH11 | 0 | 0.02 | M1 | Relapse |
| PAXJGA | CHEK2 | c.599T>C | p.(Ile157Thr) | Missense | NM_007194.4 | LP | PS3, PS4 moderate, PM1, BS1 | BMRS | NA | 0.63 | 0.46 | 0.46 | 0.58 | F | Child | CBFB-MYH11 | 0 | 0 | Unknown | Relapse |
| PATJMY | CTC1 | c.1360del | p.(Glu454SerfsTer9) | Frameshift | NM_025099.6 | LP | PVS1, PP4 | BMSC | 0.53 | NA | NA | NA | 0.28 | M | Child | NUP98-NSD1 | NA | NA | M2 | Induction failure |

| | | | | | | | | | | | | | | | | | | | | |
|---|---|---|---|---|---|---|---|---|---|---|---|---|---|---|---|---|---|---|---|---|
| PAVIGB | CTC1 | c.277C>T | p.(Gln93Ter) | Stopgain | NM_025099.6 | LP | PVS1, PM2 supporting | BMRS | NA | 0.45 | 0.55 | NA | 0.59 | F | Infant | KMT2A-MLLT3 | 0 | 0 | Unknown | Relapse |
| PAWTWW | CUX1 | c.1754del | p.(Phe585SerfsTer11) | Frameshift | NM_001913.5 | LP | PVS1, PM2 supporting | BMRS | NA | 0.44 | 0.58 | 0.47 | 0.02 | M | AYA | KMT2A-ELL | 0 | 0 | M5 | Relapse |
| PAUSBP | DDX54 | c.2196-2A>C | Retention of intron 17/19 | Splice | NM_001111322.2 | LP | PM2 supporting, PVS1 | BMRS | NA | 0.46 | NA | 0.49 | 0.9 | M | Child | KMT2A-AFDN | 0 | 0 | M5 | Relapse |
| PAWSJC | DHX34 | c.1621C>T | p.(Arg541Ter) | Stopgain | NM_014681.6 | LP | PVS1, PM2 supporting | BMRS | NA | 0.66 | 0.45 | 0.41 | 0.48 | F | AYA | CBFB-MYH11 | 0 | 0 | Unknown | Relapse |
| PAWSJC | DHX34 | c.2463_2469del | p.(Ser821ArgfsTer50) | Frameshift | NM_014681.6 | LP | PVS1, PM2 supporting | BMRS | NA | 0.38 | 0.32 | 0.36 | 0.15 | F | AYA | CBFB-MYH11 | 0 | 0 | Unknown | Relapse |
| PAUVBS | DNAH5 | c.9502C>T | p.(Arg3168Ter) | Stopgain | NM_001369.3 | LP | PVS1, PM2 supporting | BMRS | NA | 0.39 | 0.53 | NA | 0* | F | Child | KMT2A-MLLT1 | 0.03 | 4.7 | M1 | Relapse |
| PAVDRG | DNAH5 | :c.328A>T | p.(Lys110Ter) | Stopgain | NM_001369.3 | LP | PVS1, PM2 supporting | BMRS | NA | 0.5 | 0.3 | NA | 0* | F | Child | DEK_NUP214 | 0.03 | 0 | Unknown | Censored |
| PAWBSJ | DNAH5 | c.11725C>T | p.(Arg3909Ter) | Stopgain | NM_001369.3 | LP | PVS1, PP4 | BMRS | NA | 0.52 | 0.34 | 0.58 | 0* | M | Child | RUNX1-RUNX1T1 | 0.04 | 0 | Unknown | Relapse |
| PAXELP | DNAH5 | c.13194_13197del | p.(Asp4398GlufsTer16) | Frameshift | NM_001369.3 | P | PVS1, PM3, PP4 | BMRS | NA | 0.57 | 0.57 | 0.45 | 0* | M | AYA | RUNX1-RUNX1T1 | 0.04 | 1.4 | Unknown | Relapse |
| PAVCNG | DNAH9 | c.9421C>T | p.(Gln3141Ter) | Stopgain | NM_001372.4 | LP | PVS1, PM2 supporting | BMRS | NA | 0.45 | 0.44 | NA | 0* | M | AYA | CEBPA | 0 | 0 | M2 | Relapse |
| PAVCNU | DNAH9 | c.308del | p.(Phe103SerfsTer31) | Frameshift | NM_001372.4 | LP | PVS1, PM3 supporting | BMRS | NA | 0.48 | 0.44 | NA | 0* | F | Child | NUP98-HOXA13 | 0 | 0 | M2 | Death |
| PARBTV | DNMT3A | c.2645G>A | p.(Arg882His) | Missense | NM_022552.5 | P | PS4 very strong, PS3, PP3 | BMSC | 0.54 | NA | NA | NA | 0.49 | M | AYA | None | NA | NA | M1 | Induction failure |
| PAWMKP | ERBB2 | c.30dup | p.(Leu11AlafsTer99) | Frameshift | NM_004448.4 | LP | PM2 supporting, PVS1 | BMRS | NA | 0.59 | NA | 0.44 | 0* | M | Child | RUNX1-RUNX1T1 | 0 | 0 | Unknown | Relapse |
| PAWAMB | FANCA | c.1615del | p.(Asp539ThrfsTer66) | Frameshift | NM_000135.4 | LP | PVS1, PM2 supporting | BMRS | NA | 0.62 | 0.44 | 0.49 | 0.34 | F | Infant | NUP98-NSD1 | 0 | 0 | Unknown | Relapse |

| | | | | | | | | | | | | | | | | | | | | |
|---|---|---|---|---|---|---|---|---|---|---|---|---|---|---|---|---|---|---|---|---|
| PAVBUX | FANCD2 | c.982C>T | p.(Arg328Ter) | Stopgain | NM_033084.6 | LP | PVS1, PM2 supporting | BMRS | NA | 0.46 | 0.41 | NA | 0.38 | M | AYA | KMT2A PTD | KMT2A PTD in tumor WGS and absent in remission WGS | 0 | M4 | Relapse |
| PAXELP | FANCM | c.5791C>T | p.(Arg1931Ter) | Stopgain | NM_020937.4 | LP | PVS1 strong, PS3-supporting, PM1 | BMRS | NA | 0.34 | 0.43 | 0.51 | 0* | M | AYA | RUNX1-RUNX1T1 | 0.04 | 1.4 | Unknown | Relapse |
| PAUUKZ | FGFR3 | c.445+2T>G | Retention of intron 4/17 | Splice | NM_000142.5 | LP | PVS1 (RNA), PM2 supporting | BMRS | NA | 0.6 | 0.59 | 0.48 | NA | M | Infant | KMT2A-MLLT3 | 0 | 0 | Unknown | Relapse |
| PAUWKY | GATA2 | c.241G>T | p.(Gly81Ter) | Stopgain | NM-032638.5 | LP | PVS1, PM2 supporting | BMRS | NA | 0.43 | 0.61 | NA | 0.70 | M | Child | None | 0 | NA | Unknown | Death |
| PAVEAF | GBA1 | c.1483G>C | p.(Ala495Pro) | Missense | NM_001005742.3 | P | PS3, PS4, PM3 | BMRS | NA | 0.56 | 0.22 | NA | 0* | M | Child | NUP98-NSD1 | 0 | 7.4 | Unknown | Relapse |
| PAVGKS | GBA1 | c.1226A>G | p.(Asn409Ser) | Missense | NM_001005742.3 | LP | PS3 moderate, PS4, PM1, PM3, PP1, PP2 | BMRS | NA | 0.47 | 0.43 | 0.56 | 0* | F | AYA | KMT2A-ELL | 0 (FLT3-ITD) | 0 | NOS | Relapse |
| PAVKKI | GBA1 | c.882T>G | p.(His294Gln) | Missense | NM_001005742.3 | LP | PM3 Strong, PS4 Moderate | BMRS | NA | 0.5 | 0.24 | NA | 0* | F | Child | NUP98-NSD1 | 0.04 | 18 | M0 | Relapse |
| PAWPKR | HCLS1 | c.1191T>G | p.(Tyr397Ter) | Stopgain | NM_005335.6 | LP | PVS1, PM2 supporting | BMRS | NA | 0.58 | 0.72 | 0.68 | NA | F | Child | KMT2A-MLLT3 | 0 | 0 | Unknown | Relapse |
| PAVRJP | INPP5D | c.349+2T>G | Retention of intron 3/26 | Splice | NM_005541.5 | LP | PVS1 (RNA), PP3 | BMRS | NA | 0.33 | NA | 0.33 | 0.75 | M | AYA | KMT2A-MLLT10 | 0 | 0 | M5 | Relapse |
| PAUMCV | MITF | c.1273G>A | p.(Glu425Lys) | Missense | NM_001354604.2 | LP | PS3 moderate, PS4 moderate, PM1, PP1, BP4 | BMRS | NA | 0.47 | 0.54 | NA | NA | F | Infant | None | del(13q) present in tumor WGS and absent in remission WGS | 0 | M6 | Censored |
| PAVMEH | MITF | c.1273G>A | p.(Glu425Lys) | Missense | NM_001354604.2 | LP | PS3 moderate, PS4 moderate, PM1, PP1, BP4 | BMRS | NA | 0.38 | 0.21 | 0.54 | 0* | F | Child | CEBPA | 0 | 0 | Unknown | Censored |

| | | | | | | | | | | | | | | | | | | | | |
|---|---|---|---|---|---|---|---|---|---|---|---|---|---|---|---|---|---|---|---|---|
| PAWPKR | MITF | c.1273G>A | p.(Glu425Lys) | Missense | NM_001354604.2 | LP | PS3 moderate, PS4 moderate, PM1, PP1, BP4 | BMRS | NA | 0.59 | 0.44 | 0.63 | NA | F | Child | KMT2A-MLLT3 | 0 | 0 | Unknown | Relapse |
| PAUVWV | MRE11 | c.1714C>T | p.(Arg572Ter) | Stopgain | NM_005591.4 | P | PVS1, PS4 | BMRS | NA | 0.58 | 0.4 | NA | 0* | F | Infant | KMT2A-MYO1F | 0 | 0 | Unknown | Censored |
| PAUMGZ | MUTYH | c.1187G>A | p.(Gly396Asp) | Missense | NM_001128425.2 | P | PS3, PS4, PP3 | BMRS | NA | 0.6 | 0.53 | NA | 0* | F | Child | NPM1 | 0 | 0 | M1 | Censored |
| PAUMUZ | MUTYH | c.1187G>A | p.(Gly396Asp) | Missense | NM_001128425.2 | P | PS3, PS4, PP3 | BMRS | NA | 0.49 | 0.25 | 0.41 | 0.00 | F | Child | MYB_CLINT1 | 0 | 0 | Unknown | Relapse |
| PAUNVK | MUTYH | c.1187G>A | p.(Gly396Asp) | Missense | NM_001128425.2 | P | PS3, PS4, PP3 | BMRS | NA | 0.5 | 0.64 | NA | 0.5 | M | Child | RUNX1-RUNX1T1 | 0 | 0 | Unknown | Censored |
| PAUVWV | MUTYH | c.536A>G | p.(Tyr179Cys) | Missense | NM_001128425.2 | P | PS3, PS4, PP3 | BMRS | NA | 0.58 | 0.52 | NA | 0.63 | F | Infant | KMT2A-MYO1F | 0 | 0 | Unknown | Censored |
| PAWUAE | MUTYH | c.1187G>A | p.(Gly396Asp) | Missense | NM_001128425.2 | P | PS3, PS4, PP3 | BMRS | NA | 0.54 | 0.49 | 0.43 | 0.52 | M | Infant | KMT2A-MLLT10 | 0 | 0 | Unknown | Relapse |
| PAVHLI | MYO5A | c.2416C>T | p.(Arg806Ter) | Stopgain | NM_000259.3 | LP | PVS1, PM2 supporting | BMRS | NA | 0.61 | NA | 0.4 | NA | F | AYA | CBFB-MYH11 | 0 | 0 | Unknown | Relapse |
| PAVBYN | NFKBIE | c.137del | p.(Ile46ThrfsTer3) | Frameshift | ENST00000275015.5 | LP | PVS1, PM2 supporting | BMRS | NA | 0.5 | NA | 0.48 | 0* | F | AYA | CEBPA | 0 | 0 | Unknown | Relapse |
| PAULCA | NTHL1 | c.835C>T | p.(Gln279Ter) | Stopgain | NM_002528.7 | P | PVS1 strong, PM3 supporting, PS3 | BMRS | NA | 0.56 | 0.63 | NA | 0.45 | F | Infant | None | Unable to identify somatic driver | 8.9 | M6 | Death without remission |
| PAXETC | PALB2 | c.3113G>A | p.(Trp1038Ter) | Stopgain | NM_024675.4 | P | PVS1, PS4 moderate, PP1 | BMRS | NA | 0.39 | 0.42 | 0.36 | 0.11 | M | AYA | KMT2A_MYOCD | +8 in tumor WGS and absent in remission WGS | 37 | M5 | Relapse |
| PAUNSV | PRF1 | c.50del | p.(Leu17ArgfsTer34) | Frameshift | NM_005041.6 | P | PVS1, PS3, PS4 | BMRS | NA | 0.36 | NA | 0.38 | 0.12 | M | Child | CEBPA | 0 | 0 | Unknown | Relapse |
| PAVFDW | PRF1 | c.50del | p.(Leu17ArgfsTer34) | Frameshift | NM_005041.6 | P | PVS1, PS3, PS4 | BMRS | NA | 0.49 | 0.43 | NA | 0.75 | F | AYA | NPM1 | 0 | 0 | M2 | Relapse |

| | | | | | | | | | | | | | | | | | | | |
|---|---|---|---|---|---|---|---|---|---|---|---|---|---|---|---|---|---|---|---|
| PAVHCV | PRF1 | c.50del | p.(Leu17ArgfsTer34) | Frameshift | NM_005041.6 | P | PVS1, PS3, PS4 | BMRS | NA | 0.53 | 0.61 | NA | 0.12 | M | Child | CBFB-MYH11 | 0 | 0 | Unknown | Censored |
| PAXELP | PRF1 | c.666C>A | p.(His222Gln) | Missense | NM_005041.6 | P | PS4, PS3, PM1, PP3 | BMRS | NA | 0.5 | 0.67 | 0.55 | 0.67 | M | AYA | RUNX1-RUNX1T1 | 0.04 | 1.4 | Unknown | Relapse |
| PAUZVP | PRPF8 | c.605_606del | p.(Pro202ArgfsTer7) | Frameshift | NM_006445.4 | LP | PVS1, PM2 supporting | BMRS | NA | 0.5 | 0.55 | 0.38 | 0.22 | M | AYA | NUP214-ABL1 | XXYY present in tumor WGS and absent in remission WGS | 5 | Unknown | Relapse |
| PAXMLI | PRPF8 | c.6991del | p.(Glu2331ArgfsTer28) | Frameshift | NM_006445.4 | LP | PVS1 strong, PM2 supporting, PP1 | BMRS | NA | 0.39 | NA | 0.44 | 0.44 | M | AYA | KMT2A-MLLT10 | 0 | 0 | M5 | Relapse |
| PAXBAZ | RAD50 | c.832C>T | p.(Arg278Ter) | Stopgain | NM_005732.4 | LP | PVS1, PP4 | BMRS | NA | 0.45 | 0.58 | 0.63 | 0.56 | F | Child | TEC-MLLT10 | 0 | 0 | M7 | Relapse |
| PAVTDU | RAD51C | c.577C>T | p.(Arg193Ter) | Stopgain | NM-058216.3 | P | PVS1, PS4 | BMRS | NA | 0.63 | NA | 0.51 | 0* | F | Child | None | 0 | 0 | M2 | Relapse |
| PAVLJH | RASA2 | c.874C>T | p.(Gln292Ter) | Stopgain | NM_006506.5 | LP | PVS1, PM2 supporting | BMRS | NA | 0.43 | 0.43 | 0.5 | 0.23 | F | AYA | CBFB-MYH11 | 0 | 0 | Unknown | Relapse |
| PAWCJK | RECQL4 | c.3072del | p.(Val1026CysfsTer18) | Frameshift | NM_004260.4 | P | PVS1, PM2 supporting, PM3 supporting | BMRS | NA | 0.61 | 0.33 | 0.49 | 0.29 | F | AYA | NPM1 | 0 | 0 | Unknown | Relapse |
| PAUUKK | RECQL5 | c.2791A>T | p.(Lys931Ter) | Stopgain | NM_004259.7 | LP | PVS1 (RNA), PM2 supporting | BMRS | NA | 0.38 | 0.62 | NA | 0.43 | F | Infant | KAT6A-CREBBP | 0 | 0 | M5 | Relapse |
| PAVCNG | ROS1 | c.4190G>A | p.(Trp1397Ter) | Stopgain | NM_002944.3 | LP | PVS1, PM2 supporting | BMRS | NA | 0.38 | 0.46 | NA | 0* | M | AYA | CEBPA | 0 | 0 | M2 | Relapse |
| PAXIFA | ROS1 | c.350del | p.(Leu117TyrfsTer11) | Frameshift | NM_002944.3 | LP | PVS1, PM2 supporting | BMRS | NA | 0.45 | 0.56 | 0.54 | NA | M | Child | KMT2A-MLLT10 | 0 | 0.3 | Unknown | Induction failure |
| PAVGBH | RPS27A | c.-17-98T>A | Retention of intron 1 (upstream of start) | Splice | NM_001135592.2 | LP | PVS1 (RNA), PM2 supporting | BMRS | NA | 0.3 | 0.38 | NA | 0.96 | M | Infant | None | 0 | 0 | M5 | Censored |

| | | | | | | | | | | | | | | | | | | | | |
|---|---|---|---|---|---|---|---|---|---|---|---|---|---|---|---|---|---|---|---|---|
| PAXJZX | SBDS | c.258+2T>C | 8 bp deletion at the beginning of exon 2 | Splice | NM-016038.4 | P | PVS1 (RNA), PS3 supporting, PM3 strong | BMRS | NA | 0.52 | NA | 0.55 | 0.22 | F | Child | KMT2A-X | 0 | 3.1 | Unknown | Relapse |
| PAXJZX | SBDS | c.258+1G>C | 8 bp deletion at the beginning of exon 2 | Splice | NM-016038.4 | P | PVS1 (RNA), PM2 supporting, PM3 supporting | BMRS | NA | 0.48 | NA | 0.47 | 0.77 | F | Child | KMT2A-X | 0 | 3.1 | Unknown | Relapse |
| PANVPB | SETDB1 | c.1214C>A | p.(Ser405Ter) | Stopgain | NM_012432.4 | LP | PVS1, PM2 supporting | BMSC | 0.6 | NA | NA | NA | 0* | M | AYA | MLLT10-PSD3 | NA | NA | M5 | Induction failure |
| PARXYR | SLX4 | c.3895_3896del | p.(Arg1299GlyfsTer35) | Frameshift | NM_032444.4 | LP | PVS1, PM2 supporting | BMSC | 0.6 | NA | NA | NA | NA | F | AYA | NUP98-NSD1 | NA | NA | M1 | Relapse |
| PAUTVU | SLX4 | c.928C>T | p.(Arg310Ter) | Stopgain | NM_032444.4 | LP | PVS1, PM2 supporting | BMRS | NA | 0.58 | 0.44 | NA | 0* | F | Infant | RBM15-MRTFA | 0 | 0 | M7 | Censored |
| PAVGAM | SMO | c.1921C>G | p.(Pro641Ala) | Missense | NM_005631.5 | LP | PS3, PP1, PP3 | BMRS | NA | 0.44 | 0.44 | 0.5 | 0* | F | Infant | KMT2A-MLLT3 | Add(17)(p13) in tumor WGS and not in remission WGS | 0 | M4 | Relapse |
| PAVVKU | SMO | c.1921C>G | p.(Pro641Ala) | Missense | NM_005631.5 | LP | PS3, PP1, PP3 | BMRS | NA | 0.49 | 0.42 | 0.27 | 0.63 | M | Infant | None | 0 (t(X;13)) | 0.2 | M7 | Relapse |
| PAVIYH | STXBP2 | c.1654G>A | p.(Gly552Ser) | Missense | NM_001272034.2 | LP | PS3, PM3, PP3 | BMRS | NA | 0.36 | 0.43 | NA | 0.46 | F | Child | KMT2A-MLLT3 | 0 | 0 | Unknown | Censored |
| PAVDVV | SUFU | c.367C>T | p.(Arg123Cys) | Missense | NM_016169.4 | LP | PP1 Strong, PS3 moderate PP3, PM2 supporting | BMRS | NA | 0.49 | 0.52 | 0.49 | 0.5 | F | AYA | KMT2A-MLLT1 | 0 | 0 | M5 | Relapse |
| PAXDEG | SYNE1 | c.23309T>A | p.(Leu7770Ter) | Stopgain | NM_182961.4 | LP | PVS1, PM2 supporting | BMRS | NA | 0.71 | 0.43 | 0.47 | 0.56 | F | AYA | None | Unable to identify somatic driver | 0 | M1 | Relapse |
| PAUMMR | TNFRSF13B | c.542C>A | p.(Ala181Glu) | Missense | NM-012452.3 | LP | PS3, PS4, PP1, BS1 | BMRS | NA | 0.43 | 0.44 | NA | 0.67* | M | AYA | NUP98-NSD1 | 0.03 | 16 | Unknown | Censored |
| PAUSFM | TNFRSF13B | c.204dup | p.(Leu69ThrfsTer12) | Frameshift | NM-012452.3 | P | PVS1, PS4, PP1 | BMRS | NA | 0.37 | 0.39 | 0.44 | 0* | M | Child | KMT2A-ELL | 0 | 0 | M2 | Censored |

| | | | | | | | | | | | | | | | | | | | | |
|---|---|---|---|---|---|---|---|---|---|---|---|---|---|---|---|---|---|---|---|---|
| PAUYAE | TNFRSF13B | c.310T>C | p.(Cys104Arg) | Missense | NM-012452.3 | LP | PS3, PS4 moderate, PP3, BS2 | BMRS | NA | 0.68 | 0.56 | NA | 0* | F | AYA | NPM1 | 0 | 0 | M4 | Censored |
| PAUYEV | TNFRSF13B | c.310T>C | p.(Cys104Arg) | Missense | NM-012452.3 | LP | PS3, PS4 moderate, PP3, BS2 | BMRS | NA | 0.48 | NA | 0.46 | 1* | M | Child | RUNX1-RUNX1T1 | 0 | 0 | Unknown | Relapse |
| PAUZSY | TNFRSF13B | c.310T>C | p.(Cys104Arg) | Missense | NM-012452.3 | LP | PS3, PS4 moderate, PP3, BS2 | BMRS | NA | 0.39 | NA | 0.49 | 0* | F | Child | None | 0 (FLT3-ITD) | 0.8 | Unknown | Relapse |
| PAWUAE | TNFRSF13B | c.310T>C | p.(Cys104Arg) | Missense | NM-012452.3 | LP | PS3, PS4 moderate, PP3, BS2 | BMRS | NA | 0.58 | 0.63 | 0.37 | 0.52* | M | Infant | KMT2A-MLLT10 | 0 | 0 | Unknown | Relapse |
| PAXJZX | TP53 | c.626G>A | p.(Arg209Gln) | Missense | NM-001276760.3 | P | PS3, PP3, PP1, PS4 supporting, PM2 supporting | BMRS | NA | 0.43 | NA | 0.75 | 0.98 | F | Child | KMT2A-X | 0 | 3.1 | Unknown | Relapse |
| PAXLGL | VPS13B | c.5120_5124del | p.(Lys1707SerfsTer9) | Frameshift | NM_017890.5 | LP | PVS1, PM2 supporting | BMRS | NA | 0.46 | NA | 0.52 | 0.27 | M | AYA | KMT2A-ELL | 0 | 0 | M2 | Relapse |
| PAVNKI | VPS45 | c.695_696del | p.(Tyr232SerfsTer16) | Frameshift | NM_007259.5 | LP | PVS1, PM2 supporting | BMRS | NA | 0.36 | 0.55 | 0.49 | 0.48 | F | AYA | CBFB-MYH11 | 0 | 0 | Unknown | Relapse |
| PAUTML | XPC | c.463C>T | p.(Arg155Ter) | Stopgain | NM_004628.5 | LP | PVS1, PM2 supporting | BMRS | NA | 0.37 | 0.44 | NA | NA | NA | NA | NA | 0.05 (NRAS G12D) | NA | NA | NA |
| PASYEJ | XRCC2 | c.96del | p.(Phe32LeufsTer30) | Frameshift | NM_005431.2 | LP | PVS1, PM2 supporting | BMSC | 0.48 | NA | NA | NA | 0.2 | F | Infant | None | NA | NA | NOS | Induction failure |
| PAXISI | ZCCHC8 | c.317+2T>C | Loss of exon 3 | Splice | NM-017612.5 | LP | PVS1 (RNA), PM2 supporting | BMRS | NA | 0.43 | NA | 0.5 | 0.67 | M | AYA | CEBPA | 0 | 0 | M4 | Relapse |

USI - unique subject identifier, ACMG - American College of Medical Genetics, AMP - Association of Molecular Pathology, P - pathogenic, LP - likely pathogenic, BMRS - Bone marrow remission sample, BMSC - Bone marrow stromal cells. VAF - variant allele frequency (%), M - male, F - female, FAB - French-American-British subtype. Age groups- Infant: 0-2 years, Child: 3-14 years, AYA: 15-40 years. *Low RNA coverage

Supplementary Table 12: Additional somatic variants in genes with P/LP variants

| USI | Gene | Transcript | Variant cDNA | Variant Protein | Diagnosis VAF |
|---|---|---|---|---|---|
| PAUUKZ | *FGFR3* | NM_001163213.2 | c.1082-510G>A | Splice | 0.07 |
| PAUZVP | *PRPF8* | NM_006445.4 | c.6929G>A | p.(Arg2310Lys) | 0.09 |
| PAUZVP | *PRPF8* | NM_006445.4 | c.867-1G>T | Splice | 0.06 |
| PAWTWW | *CUX1* | NM_001913.5 | c.1823G>A | p.(Gly608Glu) | 0.17 |

Supplementary Table 13: Genes and mode of inheritance included in cohort meta-analysis

| Gene | Inheritance |
|---|---|
| ADA | AR |
| ADA2 | AR |
| ALAS2 | XLD |
| ANKRD26 | AD |
| APOA1 | AD/AR |
| APOA2 | AD/AR |
| ARID1A | AD |
| ARID5B | Unknown |
| ATG2B | Unknown |
| ATM | AD/AR |
| BLM | AD/AR |
| BRCA1 | AD |
| BRCA2 | AD |
| BRIP1 | AR |
| BTK | XLR |
| CARD11 | AD/AR |
| CASP10 | AD |
| CBL | AD |
| CD27 | AR |
| CD40LG | XLR |
| CD70 | AR |
| CDKN2A | AD |
| CDKN2B | Unknown |
| CEBPA | AD |
| CEBPE | AD/AR |

| | |
|---|---|
| CHEK2 | AD |
| CSF3R | AD/AR |
| CST3 | AD |
| CTC1 | AR |
| CTLA4 | AD |
| CTPS1 | AR |
| CXCR4 | AD |
| DDX41 | AD |
| DIS3 | Unknown |
| DKC1 | XLR |
| DNAJC21 | AR |
| DOCK8 | AR |
| EFTUD1 | AR |
| ELANE | AD |
| ERCC4 | AR |
| ERCC6L2 | AR |
| ETV6 | AD |
| FADD | AR |
| FANCA | AR |
| FANCB | XLR |
| FANCC | AR |
| FANCD2 | AR |
| FANCE | AR |
| FANCF | AR |
| FANCG | AR |
| FANCI | AR |
| FANCL | AR |
| FANCM | AR |
| FAS | AD |
| FASLG | AD |
| FGA | AD/AR |
| G6PC3 | AR |
| GATA1 | XLR |
| GATA2 | AD |
| GATA3 | AD |
| GFI1 | AD |
| GSKIP | Unknown |

| | |
|---|---|
| GSN | AD |
| HAX1 | AR |
| IKZF1 | AD |
| ITK | AR |
| JAK2 | AD |
| KDM1A | AD |
| KLHDC8B | AR |
| LIG4 | AR/Smu |
| LYZ | AD |
| LZTR1 | AD |
| MAGT1 | XLR |
| MBD4 | AD/AR |
| MECOM | AD |
| MLH1 | AD/AR |
| MPL | AD/AR/Smu |
| MRTFA | AR |
| MSH2 | AD/AR |
| MSH6 | AD/AR |
| NAF1 | AD |
| NBN | AD/AR |
| NF1 | AD |
| NHP2 | AR |
| NOP10 | AR/AD |
| NPAT | Unknown |
| NPM1 | Smu |
| PALB2 | AR |
| PARN | AD/AR |
| PAX5 | AD |
| PGM3 | AR |
| PIK3CD | AD/AR |
| PIP4K2A | Unknown |
| PMS2 | AD |
| POT1 | AD/AR |
| PRF1 | AR |
| PTEN | AD |
| PTPN11 | AD |
| RAD51 | AD |

| | |
|---|---|
| RAD51C | AD |
| RASGRP1 | AR |
| RBBP6 | Unknown |
| RBM8A | AR |
| RECQL4 | AR |
| RPL11 | AD |
| RPL15 | AD |
| RPL18 | AD |
| RPL23 | Unknown |
| RPL26 | AD |
| RPL27 | AD |
| RPL31 | Unknown |
| RPL35 | AD |
| RPL35A | AD |
| RPL36 | Unknown |
| RPL5 | AD |
| RPS10 | AD |
| RPS17 | AD |
| RPS19 | AD |
| RPS24 | AD |
| RPS26 | AD |
| RPS27 | AD |
| RPS28 | AD |
| RPS29 | AD |
| RPS7 | AD |
| RTEL1 | AD/AR |
| RUNX1 | AD |
| SAMD9 | AD |
| SAMD9L | AD |
| SBDS | AR |
| SH2B3 | AR |
| SH2D1A | XLR |
| SLX4 | AR |
| SRP54 | AD |
| SRP72 | AD |
| STAT3 | AD |
| STXBP2 | AR |

| | |
|---|---|
| TERC | AD |
| TERF2IP | Unknown |
| TERT | AD |
| TET2 | AR/Smu |
| TINF2 | AD |
| TNFRSF13B | AD/AR |
| TNFRSF9 | AR |
| TP53 | AD |
| TTR | AD |
| UBE2T | AR |
| UNC13D | AR |
| USB1 | AR |
| USP45 | AR |
| VPS45 | AR |
| WAS | XLR |
| WRAP53 | AR |
| XRCC2 | AR |
| ZNF43 | Unknown |

#### Supplementary References:


1. McNeer NA, Philip J, Geiger H, et al. Genetic mechanisms of primary chemotherapy resistance in pediatric acute myeloid leukemia. Leukemia 2019;33(8):1934–43.

2. Li H, Durbin R. Fast and accurate short read alignment with Burrows-Wheeler transform. Bioinformatics 2009;25(14):1754–60.

3. Edmonson MN, Zhang J, Yan C, Finney RP, Meerzaman DM, Buetow KH. Bambino: a variant detector and alignment viewer for next-generation sequencing data in the SAM/BAM format. Bioinformatics 2011;27(6):865–6.

4. Huang YP, Harmon L, Gardner E, et al. bamSliceR: cross-cohort variant and allelic bias analysis for rare variants and rare diseases. bioRxiv [Internet] 2023;Available from: http://dx.doi.org/10.1101/2023.09.15.558026

5. Bolouri H, Ries R, Pardo L, et al. A B-cell developmental gene regulatory network is activated in infant AML. PLoS One 2021;16(11):e0259197.

6. Chen X, Schulz-Trieglaff O, Shaw R, et al. Manta: rapid detection of structural variants and indels for germline and cancer sequencing applications. Bioinformatics 2016;32(8):1220–2.

7. Bolouri H, Farrar JE, Triche T, et al. The molecular landscape of pediatric acute myeloid leukemia reveals

recurrent structural alterations and age-specific mutational interactions. Nat Med 2018;24(1):103–12.

8. Wu T, Hu E, Xu S, et al. clusterProfiler 4.0: A universal enrichment tool for interpreting omics data. Innovation (Camb) 2021;2(3):100141.

9. Friendly M. Visualizing Categorical Data. SAS Institute; 2000.

10. Mayakonda A, Lin D-C, Assenov Y, Plass C, Koeffler HP. Maftools: efficient and comprehensive analysis of somatic variants in cancer. Genome Res 2018;28(11):1747–56.

11. Braunstein EM, Chen H, Juarez F, et al. Germline ERBB2/HER2 coding variants are associated with increased risk of myeloproliferative neoplasms. Cancers (Basel) 2021;13(13):3246.

12. Bao R, Ng A, Sasaki M, et al. Functional common and rare ERBB2 germline variants cooperate in familial and sporadic cancer susceptibility. Cancer Prev Res (Phila) 2021;14(4):441–54.

13. Ju Y, Wang L, Ta S, et al. A germline alteration of ERBB2 increases the risk of breast cancer in Chinese Han women with a familial history of malignant tumors. Oncol Lett 2019;18(3):2885–90.

14. Perkins K, Davis J, Price S, et al. Expanding spectrum of malignancies in ALPS: A cancer predisposing syndrome? Blood 2012;120(21):2149–2149.

15. Shah S, Wu E, Rao VK, Tarrant TK. Autoimmune lymphoproliferative syndrome: an update and review of the literature. Curr Allergy Asthma Rep 2014;14(9):462.

16. Rio-Machin A, Vulliamy T, Hug N, et al. The complex genetic landscape of familial MDS and AML reveals pathogenic germline variants. Nat Commun 2020;11(1):1044.

17. Molteni E, Bono E, Gallì A, et al. Prevalence and clinical expression of germ line predisposition to myeloid neoplasms in adults with marrow hypocellularity. Blood 2023;142(7):643–57.

18. Hug N, Aitken S, Longman D, et al. A dual role for the RNA helicase DHX34 in NMD and pre-mRNA splicing and its function in hematopoietic differentiation. RNA 2022;28(9):1224–38.

19. Chaudhry MS, Gilmour KC, House IG, et al. Missense mutations in the perforin (PRF1) gene as a cause of hereditary cancer predisposition. Oncoimmunology 2016;5(7):e1179415.

20. Cannella S, Santoro A, Bruno G, et al. Germline mutations of the perforin gene are a frequent occurrence in childhood anaplastic large cell lymphoma. Cancer 2007;109(12):2566–71.

21. Grisanti K, Patadia D, Scherzer R. P195 Manifestation of hemophagocytic lymphohistiocytosis in an adolescent with a heterozygous PRF1 mutation. Ann Allergy Asthma Immunol 2016;117(5):S80.

22. Xin X, Wang N, Zhang Y. Hemophagocytic lymphohistiocytosis with a hemizygous PRF1 c.674G>A mutation. Am J Med Sci 2023;366(5):387–94.

23. McReynolds LJ, Giri N, Leathwood L, Risch MO, Carr AG, Alter BP. Risk of cancer in heterozygous relatives of patients with Fanconi anemia. Genet Med 2022;24(1):245–50.

24. Nie D, Zhang J, Wang F, et al. Fanconi anemia gene-associated germline predisposition in aplastic anemia and hematologic malignancies. Front Med 2022;16(3):459–66.

25. Tischkowitz MD, Morgan NV, Grimwade D, et al. Deletion and reduced expression of the Fanconi anemia FANCA gene in sporadic acute myeloid leukemia. Leukemia 2004;18(3):420–5.

26. Abbasi S, Rasouli M. A rare FANCA gene variation as a breast cancer susceptibility allele in an Iranian population. Mol Med Rep 2017;15(6):3983–8.

27. Xia Q, Zhao L-Y, Yan Y-D, Liao Y, Di Y-S, Xiao X-Y. A multiple primary malignancy patient with FANCA gene mutation: A case report and literature review. Front Oncol 2020;10:1199.

28. Recommendations for Preventive Care for Women with Rare Genetic Cause of Breast and Ovarian Cancer » Klinicka onkologie Journal » Linkos.cz [Internet]. [cited 2025 Feb 7];Available from: https://www.linkos.cz/english-summary/klinicka-onkologie-journal/2019-08-18-supplementum-2-en/doporucení-pro-sledovani-zen-se-vzacnejsimi-genetickymi-pricinami-nadoru-prsu-a-1/

29. Chen P-C, Yin J, Yu H-W, et al. Next-generation sequencing identifies rare variants associated with Noonan syndrome. Proc Natl Acad Sci U S A 2014;111(31):11473–8.

30. Arafeh R, Qutob N, Emmanuel R, et al. Recurrent inactivating RASA2 mutations in melanoma. Nat Genet 2015;47(12):1408–10.

31. Olkinuora A, Nieminen TT, Douglas S, et al. Identification of DHX40 as a candidate susceptibility gene for colorectal and hematological neoplasia. Leukemia 2023;37(11):2301–5.

32. Yang F, Long N, Anekpuritanang T, et al. Identification and prioritization of myeloid malignancy germline variants in a large cohort of adult patients with AML. Blood 2022;139(8):1208–21.

33. Singh M, Chaudhry P, Merchant AA. Primary cilia are present on human blood and bone marrow cells and mediate Hedgehog signaling. Exp Hematol 2016;44(12):1181–7.e2.

34. Kang MR, Kim MS, Oh JE, et al. Frameshift mutations of autophagy-related genes ATG2B, ATG5, ATG9B and ATG12 in gastric and colorectal cancers with microsatellite instability: ATG gene mutations. J Pathol 2009;217(5):702–6.

35. Yu T, Ben S, Ma L, et al. Genetic variants in autophagy-related gene ATG2B predict the prognosis of colorectal cancer patients receiving chemotherapy. Front Oncol 2022;12:876424.

36. Park JW, Kim Y, Lee S-B, et al. Autophagy inhibits cancer stemness in triple-negative breast cancer via miR-181a-mediated regulation of ATG5 and/or ATG2B. Mol Oncol 2022;16(9):1857–75.

37. Tsao AS, Wistuba I, Xia D, et al. Germline and somatic smoothened mutations in non-small-cell lung cancer are potentially responsive to hedgehog inhibitor vismodegib. JCO Precis Oncol 2017;1(1):1–10.

38. Isidori F, Bozzarelli I, Ferrari S, et al. RASAL1 and ROS1 gene variants in hereditary BReast CAncer. Cancers (Basel) 2020;12(9):2539.

39. de Garibay GR, Díaz A, Gaviña B, et al. Low prevalence of SLX4 loss-of-function mutations in non-BRCA1/2 breast and/or ovarian cancer families. Eur J Hum Genet 2013;21(8):883–6.

40. Shah S, Kim Y, Ostrovnaya I, et al. Assessment of SLX4 Mutations in Hereditary Breast Cancers. PLoS One 2013;8(6):e66961.

41. Neidhardt G, Hauke J, Ramser J, et al. Association between loss-of-function mutations within the FANCM gene and early-onset familial breast cancer. JAMA Oncol 2017;3(9):1245–8.

42. Figlioli G, Bogliolo M, Catucci I, et al. The FANCM:p.Arg658* truncating variant is associated with risk of triple-negative breast cancer. NPJ Breast Cancer 2019;5(1):38.

43. Tavera-Tapia A, de la Hoya M, Calvete O, et al. RECQL5: Another DNA helicase potentially involved in hereditary breast cancer susceptibility: TAVERA-TAPIAet al. Hum Mutat 2019;40(5):566–77.

44. He Y-J, Qiao Z-Y, Gao B, Zhang X-H, Wen Y-Y. Association between RECQL5 genetic polymorphisms and susceptibility to breast cancer. Tumour Biol 2014;35(12):12201–4.

45. Shen W, Kerr CM, Przychozen B, et al. Impact of germline CTC1 alterations on telomere length in

acquired bone marrow failure. Br J Haematol 2019;185(5):935–9.

46. Fan C, Zhang J, Ouyang T, et al. RAD50 germline mutations are associated with poor survival in BRCA1/2-negative breast cancer patients. Int J Cancer 2018;143(8):1935–42.

47. Stastna B, Dolezalova T, Matejkova K, et al. Germline pathogenic variants in the MRE11, RAD50, and NBN (MRN) genes in cancer predisposition: A systematic review and meta-analysis. Int J Cancer 2024;155(9):1604–15.

48. Kluźniak W, Wokołorczyk D, Rusak B, et al. Inherited variants in XRCC2 and the risk of breast cancer. Breast Cancer Res Treat 2019;178(3):657–63.

49. Win AK, Cleary SP, Dowty JG, et al. Cancer risks for monoallelic MUTYH mutation carriers with a family history of colorectal cancer. Int J Cancer 2011;129(9):2256–62.

50. Win AK, Reece JC, Dowty JG, et al. Risk of extracolonic cancers for people with biallelic and monoallelic mutations in MUTYH. Int J Cancer 2016;139(7):1557–63.

51. Zhu M, Chen X, Zhang H, et al. AluYb8 insertion in the MUTYH gene and risk of early-onset breast and gastric cancers in the Chinese population. Asian Pac J Cancer Prev 2011;12(6):1451–5.

52. Rennert G, Lejbkowicz F, Cohen I, Pinchev M, Rennert HS, Barnett-Griness O. MutYH mutation carriers have increased breast cancer risk. Cancer 2012;118(8):1989–93.

53. Barreiro RAS, Sabbaga J, Rossi BM, et al. Monoallelic deleterious MUTYH germline variants as a driver for tumorigenesis. J Pathol 2022;256(2):214–22.

54. Li N, Zethoven M, McInerny S, et al. Evaluation of the association of heterozygous germline variants in NTHL1 with breast cancer predisposition: an international multi-center study of 47,180 subjects. NPJ Breast Cancer 2021;7(1):52.

55. Maciaszek JL, Oak N, Chen W, et al. Enrichment of heterozygous germline RECQL4 loss-of-function variants in pediatric osteosarcoma. Cold Spring Harb Mol Case Stud 2019;5(5):a004218.

56. Martin-Giacalone BA, Rideau T-T, Scheurer ME, Lupo PJ, Wang LL. Cancer risk among RECQL4 heterozygotes. Cancer Genet 2022;262-263:107–10.

57. Francisco G, Menezes PR, Eluf-Neto J, Chammas R. XPC polymorphisms play a role in tissue-specific carcinogenesis: a meta-analysis. Eur J Hum Genet 2008;16(6):724–34.

58. Zhou H, Saliba J, Sandusky GE, Sears CR. XPC protects against smoking- and carcinogen-induced lung adenocarcinoma. Carcinogenesis 2019;40(3):403–11.

59. Strom SS, Estey E, Outschoorn UM, Garcia-Manero G. Acute myeloid leukemia outcome: role of nucleotide excision repair polymorphisms in intermediate risk patients. Leuk Lymphoma 2010;51(4):598–605.

60. Liu C, Zhao L, Zhao J, Xu Q, Song Y, Wang H. Decreased ADAMTS-13 level is related to inflammation factors and risk stratification of acute lymphoblastic leukemia patients. Medicine (Baltimore) 2017;96(7):e6136.

61. Liu C, Han M, Zhao L, et al. ADAMTS-13 activity reduction in plasma of acute myeloid leukemia predicts poor prognosis after bone marrow transplantation. Hematology 2019;24(1):129–33.

62. Taddei TH, Kacena KA, Yang M, et al. The underrecognized progressive nature of N370S Gaucher disease and assessment of cancer risk in 403 patients. Am J Hematol 2009;84(4):208–14.

63. Qiu Z, Wang X, Yang Z, et al. GBA1-dependent membrane glucosylceramide reprogramming promotes

liver cancer metastasis via activation of the Wnt/β-catenin signalling pathway. Cell Death Dis 2022;13(5):508.

64. Aly M, Ramdzan ZM, Nagata Y, et al. Distinct clinical and biological implications of CUX1 in myeloid neoplasms. Blood Adv 2019;3(14):2164–78.

65. Oppermann H, Marcos-Grañeda E, Weiss LA, et al. CUX1-related neurodevelopmental disorder: deep insights into phenotype-genotype spectrum and underlying pathology. Eur J Hum Genet 2023;31(11):1251–60.

66. Huang D, Nagata Y, Grossmann V, et al. BRCC3 mutations in myeloid neoplasms. Haematologica 2015;100(8):1051–7.

67. Houzelstein D, Bullock SL, Lynch DE, Grigorieva EF, Wilson VA, Beddington RSP. Growth and early postimplantation defects in mice deficient for the bromodomain-containing protein Brd4. Mol Cell Biol 2002;22(11):3794–802.

68. Moura S, Hartl I, Brumovska V, et al. Exploring FGFR3 mutations in the male germline: Implications for clonal germline expansions and paternal age-related dysplasias. Genome Biol Evol 2024;16(2):evae015.

69. Helgason CD, Damen JE, Rosten P, et al. Targeted disruption of SHIP leads to hemopoietic perturbations, lung pathology, and a shortened life span. Genes Dev 1998;12(11):1610–20.

70. Galvin RT, Zheng C, Fitzpatrick G, et al. MYO5A::FGFR1 represents a novel fusion event in pediatric low-grade glioma. Neurooncol Adv 2023;5(1):vdad017.

71. Alves CP, Moraes MH, Sousa JF, et al. Myosin-Va contributes to manifestation of malignant-related properties in melanoma cells. J Invest Dermatol 2013;133(12):2809–12.

72. Bonato A, Chakraborty S, Bomben R, et al. NFKBIE mutations are selected by the tumor microenvironment and contribute to immune escape in chronic lymphocytic leukemia. Leukemia 2024;38(7):1511–21.

73. Lazaro-Camp VJ, Salari K, Meng X, Yang S. SETDB1 in cancer: overexpression and its therapeutic implications. Am J Cancer Res 2021;11(5):1803–27.
